# Large language model-augmented implicit surgical video review

**DOI:** 10.64898/2026.08.25.26361071

**Authors:** Zheyuan Zhang, Muhammad Ibtsaam Qadir, Rushil Ramchand, Medha Belwadi, Ryan Patrick Ball, Kali Konstantinopoulos, Evan Mark Abbey, Kathleen T. Ernsberger, Michael J. Guzman, Samantha Hendren, Bryan K. Holcomb, Bruce W. Robb, Trista Stankowski, Joshua A. Waters, Dimitrios Stefanidis, Karl Y. Bilimoria, Sanjay Mohanty, Fiona R. Kolbinger

## Abstract

Surgical video interpretation is a promising medical artificial intelligence application. However, no existing video annotation method preserves the spatiotemporal complexity of surgeon reasoning. Here we show that verbal reasoning and visual attention can be converted into structured, machine-actionable records of intraoperative behaviours. Our method decomposes transcribed verbal commentary into video-anchored semantic feedback chunks, which are classified via a large language model, with spatial grounding to surgical scenes via eyegaze or cursor tracking. We demonstrate method validity and scalability on structured and unstructured annotation tasks. For quality feedback on full-length colorectal procedures, the method reached near-human fidelity for chunking (cosine similarity: 0.95±0.01) and semantic classification across observations (Cohen’s κ: 0.71±0.07) and evaluative triggers (Cohen’s κ: 0.67±0.14), with excellent usability ratings. For structured critical view of safety assessment in laparoscopic cholecystectomy, implicit annotation yielded excellent agreement with explicit reviewer ratings (Cohen’s κ: 0.83, 0.49 and 0.81 across three criteria). We anticipate this method will advance surgical data science by enabling scalable construction of meaningfully annotated surgical video datasets.

## Introduction

Surgical procedures are a cornerstone of modern medicine and offer effective treatment for a range of acute and chronic conditions. Worldwide, over 300 million inpatient procedures are performed annually^1^, with every American adult, on average, undergoing 9.2 surgeries over their lifetime.^2^ Intraoperative surgical processes are related to patient outcomes, including 30-day morbidity after bariatric and colorectal surgery^3–5^ and long-term functional recovery after prostatectomy.^6^ Surgical departments have increasingly adopted routine documentation of video recordings of minimally invasive surgical procedures.^7^ These videos provide an opportunity to capture anatomical challenges, procedural efficiency, and technique throughout a procedure, and to support surgical assessment and coaching.

The increasing availability of computational methods for the interpretation of visual data since the mid-2010s has created opportunities to operationalise surgical video data and fostered the research field of surgical data science.^8,9^ Compared to other medical imaging data, surgical video data present distinct challenges.^8^ First, compared to static data like CT or X-ray imaging, video data comprise a temporal and a spatial component.^10^ Second, surgical procedure recordings are often several hours long, making their analysis computationally heavy.^11^ Third, surgical procedures and intraoperative behaviours are inherently unstructured and variable with regard to the sequence and duration of granular tasks like suturing or exposure and dissection of anatomic structures of interest.^12,13^ Within the same procedure type, variability can result from patient characteristics, individual surgeon preferences, or cultural and health-system differences resulting in varying availability of surgical tools or equipment.^14^ When working as a team, multiple operating surgeons can carry out visible tasks in parallel. Fourth, and critically, no video labels are routinely documented in clinical care that would be sufficiently granular on a temporal and spatial level to allow for direct training, validation, and benchmarking of computational tools on video data.^10^

While annotation strategies for surgical video data exist, they typically capture only fragmented surgeon expertise at a time, for example, temporal breakdown of a procedure into distinct phases or steps, or spatial annotation of the location of an object or anatomic structure in a single image frame isolated from the full video.^15^ Even with semi-automated methods, annotation processes often require several hours of time per hour of surgery.^16^ Non-expert crowdsourcing of annotations has shown promise for basic descriptive tasks^17–19^, yet, complex annotations to support clinically impactful surgical AI development still depend on manual or semi-manual review of video data by experienced surgeons.^20^ This has contributed to the lack of representative datasets with granular temporospatial expert annotations. Resulting computational studies reflect a limited variety of clinical tasks addressed^12^, critical model generalizability limitations, as well as remarkably poor performance of state-of-the-art artificial intelligence tools on even simple, largely descriptive surgical video understanding tasks.^21,22^

Among publicly available surgical video datasets, data scarcity manifests not only in low data volume but also in sparse annotation content. Cataract surgery and cholecystectomy alone account for 45% of publicly available surgical video datasets, and most are single-centre and modest in size. Annotations are overwhelmingly descriptive and mostly support temporal (phase, action, or step recognition) tasks through frame- or time-bound labels. Spatial labels are largely confined to isolated frames. Labels carrying clinical meaning beyond visual description are exceedingly rare, with case-level metadata and patient outcomes reported in only 9% and 3% of datasets, respectively, and all datasets relying on manual annotation.^15^ The bottleneck is therefore not a shortage of clinically important questions but the cost of the expert effort needed to convert operative footage into meaningful, spatiotemporally grounded labels. Overall, data scarcity presents a central barrier to the clinical translation of artificial intelligence in surgery.

In this context, this study was designed to test a novel method for scalable annotation of surgical video, using a large language model to classify verbal expert feedback with spatial grounding to the surgical scene via eye gaze or cursor tracking. The overarching goal was to establish the validity, scalability, and usability of this approach across two contrasting tasks: open-ended peer feedback on full-length colorectal procedures, and structured critical view of safety (CVS) assessment in laparoscopic cholecystectomy.

## Results

### Method design and working principle

To facilitate scalable surgical video review for diverse tasks, we here present a method that captures surgeon cognition, tied to surgical video, as it naturally unfolds (Fig. 1a). While reviewing a recording, an expert narrates their assessment as spoken commentary, and this commentary, rather than a set of predefined labels, is the primary annotation signal (Supplementary Information 1). Throughout the session, audio, transcript, and video are kept in temporal alignment, so that every spoken remark remains anchored to the exact moment in the procedure it refers to. By replaying the video at accelerated speed, the method allows for condensed review of longer surgery videos (Supplementary Information 2).

**Fig. 1.**
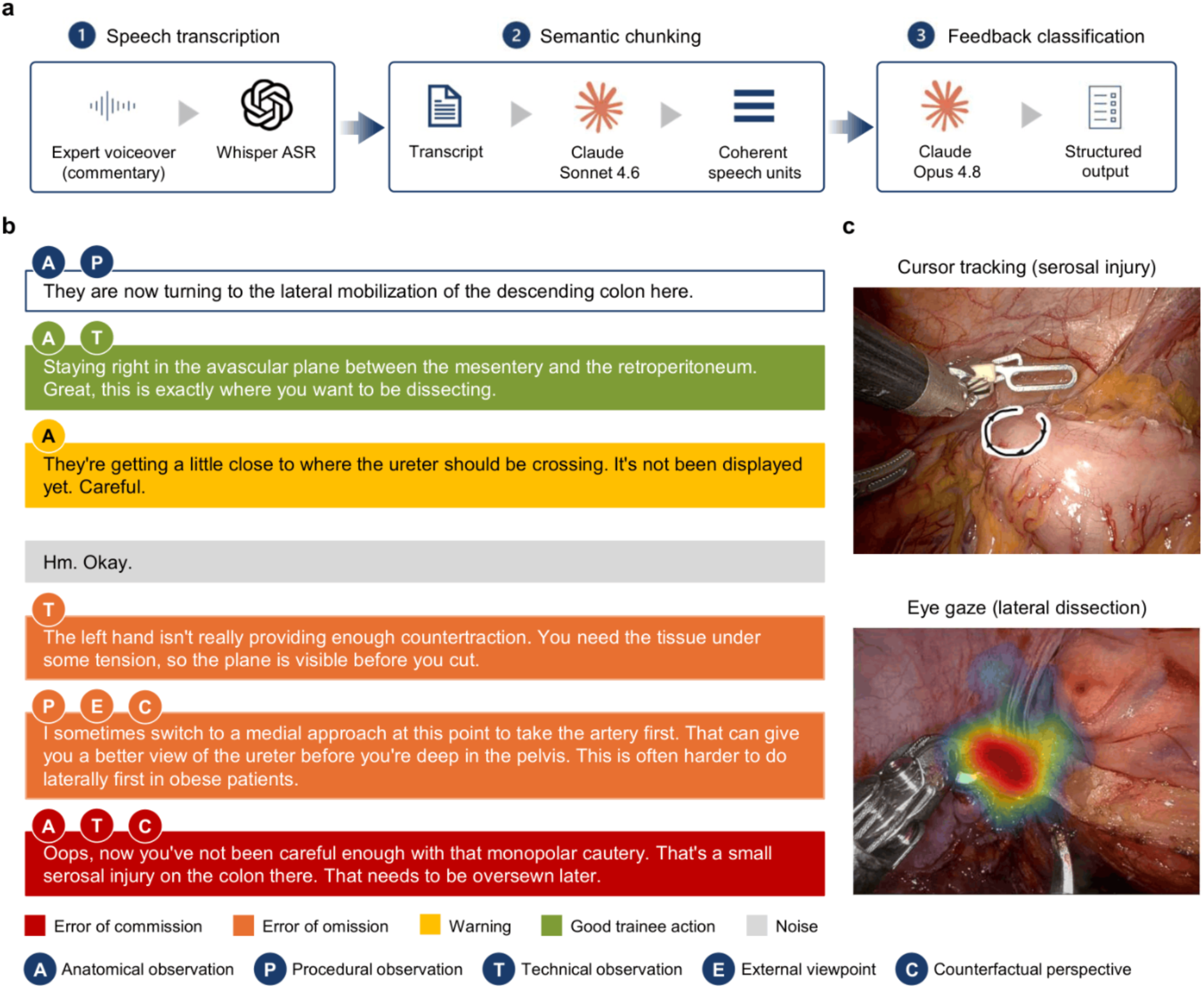
Implicit annotation framework for scalable annotation of surgical procedure recordings. a, Pipeline for automated feedback extraction from expert commentary. Experts review surgical video recordings at accelerated playback speed while providing voiceover commentary. The commentary is automatically transcribed, segmented into semantically coherent chunks, and analysed using large language models (LLMs) to generate temporally localized, structured feedback (Supplementary Information 3-9). **b,** Example of semantic chunking and feedback classification. Each text block represents a chunk corresponding to a single theme or topic. Letters denote the associated observations; a chunk may be linked to one or more observations. Box colour indicates the trigger type. Assignment of a trigger type is optional, and trigger categories are mutually exclusive. **c**, Representative examples of spatial contextualization of feedback: cursor tracking during commentary on a serosal injury (top) and eye gaze density during lateral dissection (bottom), each overlaid on the corresponding video frame. The figure displays the workflow and examples from video-based review of full procedure recordings.

The aligned transcript is then converted into structured records in two conceptual steps. First, the continuous commentary is segmented into chunks: short spans of speech, each conveying a single coherent theme, which serve as the basic unit of annotation (chunking protocol and worked examples in Supplementary Information 3-6). Second, each chunk is classified against a taxonomy that fits the prompt based on which reviewers narrated the video (classification protocol in Supplementary Information 7-9). For example, a chunk can be assigned to content and, where an evaluative stance is present, trigger categories for open-ended video-based feedback, or to a predefined criterion for structured assessment tasks (Fig. 1b). Because every chunk retains its temporal anchor, the resulting labels are localized to specific moments in the video. Spatial context can be added through the reviewer’s gaze or cursor, tying each verbal item to a region of the surgical scene (Fig. 1c): passively recorded eye gaze where dedicated hardware is available, including potential intraoperative use, or hardware-free cursor tracking for retrospective review on a standard computer screen.

In summary, this implicit review method facilitates the generation of machine-readable records linking surgical video to semantic context and spatial labels, which can subsequently be used for downstream surgical data science analyses. Of the various annotation tasks to which the method can be adapted, we validated it for two: First, unconstrained video-based peer feedback on full procedure recordings of colorectal surgery, and second, annotation of the CVS in laparoscopic cholecystectomy.

### Method validation for video-based peer feedback in colorectal surgery

We assessed the capability of implicit annotation to capture and structure complex, highly variable spatiotemporal cognitive processes by validating it on open-ended video-based feedback in colorectal surgery, in which surgeons provide peer feedback to the operating team based on the full-length procedure recording without a pre-specified taxonomy (Supplementary Information 3-9). To structure the feedback, each semantic chunk, a short span of commentary conveying a single coherent theme, was classified based on a validated surgical feedback classification system^23^ on two axes: one or more observation categories (anatomical, procedural, technical, external viewpoint or counterfactual perspective) and, where an evaluative stance was present, a single mutually exclusive feedback trigger (good trainee action, warning, error of omission or error of commission) (Fig. 1b, Fig. 2a, Supplementary Information 7-8).

**Fig. 2.**
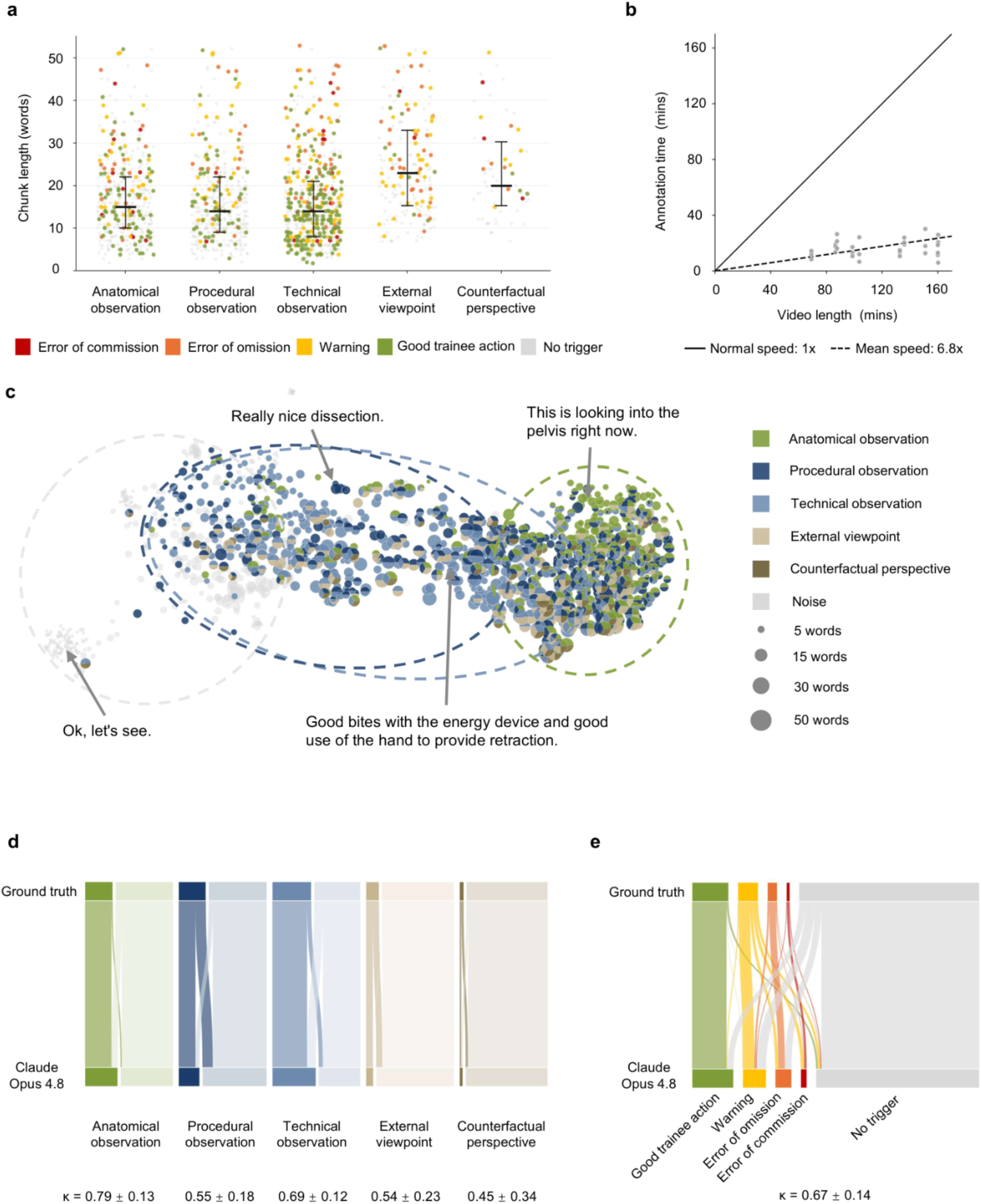
Implicit annotation of video-based peer feedback on full-length colorectal procedures. **a**, Chunk length, in words, for each observation category, represented as strip plots. Each dot indicates one chunk, coloured by trigger category. Horizontal line and whiskers represent the median and interquartile range (IQR). **b**, Annotation time versus true video length, with each dot indicating one peer’s video-based feedback. The dashed line represents the mean review speed (6.8× real time), the solid line represents hypothetical review in real-time (i.e., review duration equal to surgery duration). **c**, UMAP projection of chunk embeddings extracted from reviewer commentary via implicit annotation. Each dot represents one chunk, coloured by observation category. Dot size represents chunk length in words. Representative chunks are presented. **d,e**, Vertical Sankey diagrams comparing observation (**d**) and trigger (**e**) classifications from chunk classification via the best-performing end-to-end language model (Claude Opus 4.8, bottom) with arbitrated human ground truth (top). Ribbon width is proportional to chunk count; present assignments are shown in full colour and absent assignments as a light tint of the same colour (**d**, **e**). Cohen’s κ is reported for each category as the mean ± s.d, i.e., aggregated transcript-wise means, weighted by the number of ground-truth chunks per transcript.

The implicit annotation method decomposed the 30 video-based feedback transcripts into a total of 1,399 semantic chunks (42 [IQR 28-54] chunks per transcript). Reviewers required a mean of 8.8 ± 3.6 minutes to provide implicit peer feedback per one hour of procedure video (Fig. 2b). These chunks were then labelled independently by human annotators to establish classification ground truth, against which the model’s labels were later compared. Of the 1,399 chunks, 1,023 (73%) carried codable surgical content and 376 (27%) were non-informative (noise). Technical observations were most frequent (n = 640 chunks), followed by anatomical (n = 455) and procedural (n = 446), whereas external-viewpoint (n = 214) and counterfactual (n = 66) observations were comparatively rare. Chunk length varied by observation category: anatomical, procedural and technical observations were typically brief at mean lengths of 15 [IQR 10-22], 14 [IQR 9-22] and 14 [IQR 8-21] words, respectively, whereas external-viewpoint (23 [IQR 15-33] words) and counterfactual perspective chunks (20 [IQR 15-30] words) were longer, consistent with their more discursive, reasoning-based phrasing (Fig. 2a).

An unsupervised uniform manifold approximation and projection (UMAP) of these chunk embeddings, coloured by their arbitrated human labels, separated the non-informative (noise) chunks from codable feedback and revealed distinct semantic concepts related to chunk content categories (overall silhouette score: 0.18; Fig. 2c). Specifically, anatomical observations formed a coherent cluster, whereas procedural and technical observations frequently co-occurred within the same chunk (46% of procedural chunks were also labelled technical) and possessed semantically overlapping patterns. External-viewpoint and counterfactual chunks co-occurred with other observations in 90% and 94%, respectively, and were conceptually similar to other categories (Fig. 2c).

We further validated the automated chunking and classification against human-annotated ground truths. Producing the human chunking ground truth was itself labour-intensive, requiring a mean of 7.7 ± 6.4 minutes of manual input per transcript. Automated chunking closely reproduced this human segmentation: across the 30 transcripts, the mean cosine similarity between the model’s chunks and the arbitrated human ground truth was 0.951 ± 0.013 (aggregated transcript-wise means, weighted by the number of ground-truth chunks per transcript). This was comparable to the agreement between two independent human annotators segmenting the same transcripts into chunks, computed identically (cosine similarity: 0.963 ± 0.013, Supplementary Information 10), indicating that the model’s chunking is essentially as close to the arbitrated ground truth as one human annotator is to another.

Content agreement between implicit annotation and human-annotated ground truths was highest for concrete, visually grounded categories (anatomical: κ = 0.79 ± 0.13; technical: κ = 0.69 ± 0.12) and lower for rarer categories or those defined by reviewer stance rather than on-screen content (procedural: κ = 0.55 ± 0.18; external viewpoint: κ = 0.54 ± 0.23; counterfactual perspective: κ = 0.45 ± 0.34). The overall content agreement of the implicit annotation and the consensus-annotated ground truth was substantial (κ = 0.71 ± 0.07) (Fig. 2d). Trigger classification likewise agreed substantially with the manually annotated ground truth (κ = 0.67 ± 0.14), with disagreement arising both between the no-trigger category and the evaluative triggers and between adjacent trigger categories such as error of omission and warning (Fig. 2e).

### Implicit critical view of safety annotation in laparoscopic cholecystectomy

We further validated implicit annotation on CVS assessment in laparoscopic cholecystectomy (Supplementary Information 14-16). The CVS is a safety checkpoint in laparoscopic cholecystectomy comprising three discrete criteria: isolation of the cystic duct and cystic artery (C1), clearance of the hepatocystic triangle (C2), and exposure of the cystic plate (C3) (Fig. 3a). Compared with open-ended skill feedback, it provides a more constrained and objectifiable assessment target with established ground-truth conventions.^24^

**Fig. 3.**
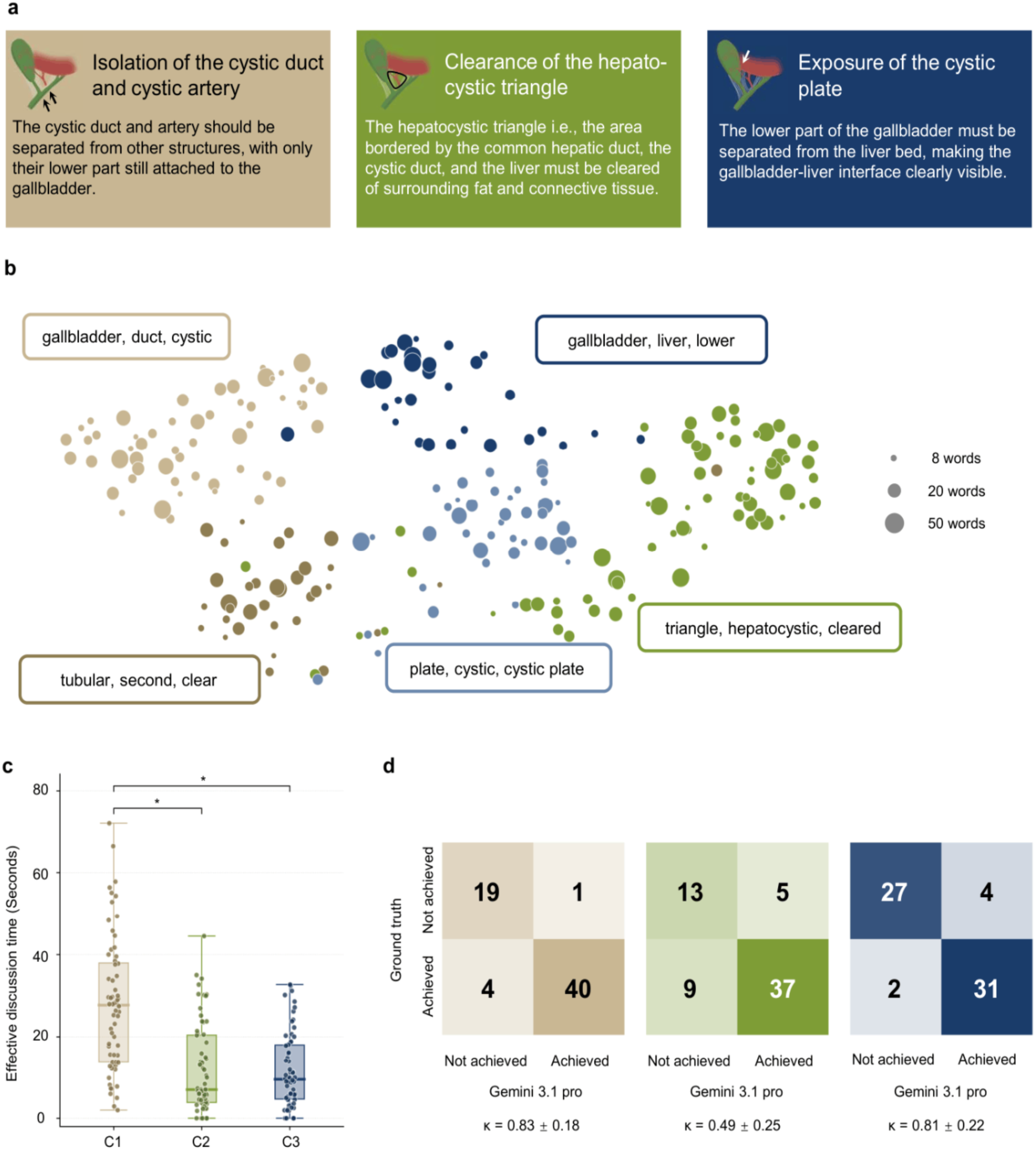
Implicit annotation of the critical view of safety (CVS) in laparoscopic cholecystectomy. a, CVS criteria. **b,** UMAP projection of chunk embeddings from reviewer commentary. Each dot represents one chunk, colored by CVS criterion; dot size represents length of chunk. Most distinctive keywords per cluster are presented. **c,** Effective discussion time for each CVS criterion across clip reviews, represented as box plots. Each dot indicates one clip per criterion. * represents Wilcoxon signed-rank test *P* < 0.05. **d,** Confusion matrices comparing CVS classifications derived from implicit verbal annotations with explicit reviewer ratings for each criterion. Cohen’s κ is reported for each criterion as mean ± s.d.

Projection of the speech transcription into chunk-level semantic embedding space revealed criterion-specific organization. Chunks, classified by corresponding criterion they inform about, occupy largely distinct regions of the UMAP representation (overall silhouette score: 0.32, Fig. 3b).

Effective discussion time was significantly longer for C1 (mean time: 27.8 ± 16.3 s) than for both C2 (12.1 ± 11.0 seconds) and C3 (11.7 ± 9.1 seconds) (both *P* < 0.05), with no notable difference between C2 and C3 (*P* = 0.77, Fig. 3c), indicating a more detailed description of cystic duct and artery isolation. When compared against the explicit per-criterion ratings provided by reviewers, LLM-augmented implicit labels were reproduced with high agreement for C1 (Cohen’s κ = 0.83) and C3 (κ = 0.81), and moderate agreement for C2 (κ = 0.49) (Fig. 3d). These findings highlight the sufficiency of semantic information in unstructured commentary to distinguish among CVS criteria.

### Spatial contextualization of implicit labels

We assessed two strategies to spatially contextualize implicit labels: eye tracking (Fig. 4a) and cursor tracking (Fig. 4b). Eye tracking provides denser, passively acquired spatial data and is best suited to settings where dedicated hardware and calibration are feasible, including potential intraoperative use. Cursor-based annotation requires no specialised hardware or calibration beyond a standard mouse or trackpad, making it more readily scalable to large-volume postoperative peer review across institutions, at the cost of lower temporal density and dependence on active reviewer engagement.

**Fig. 4.**
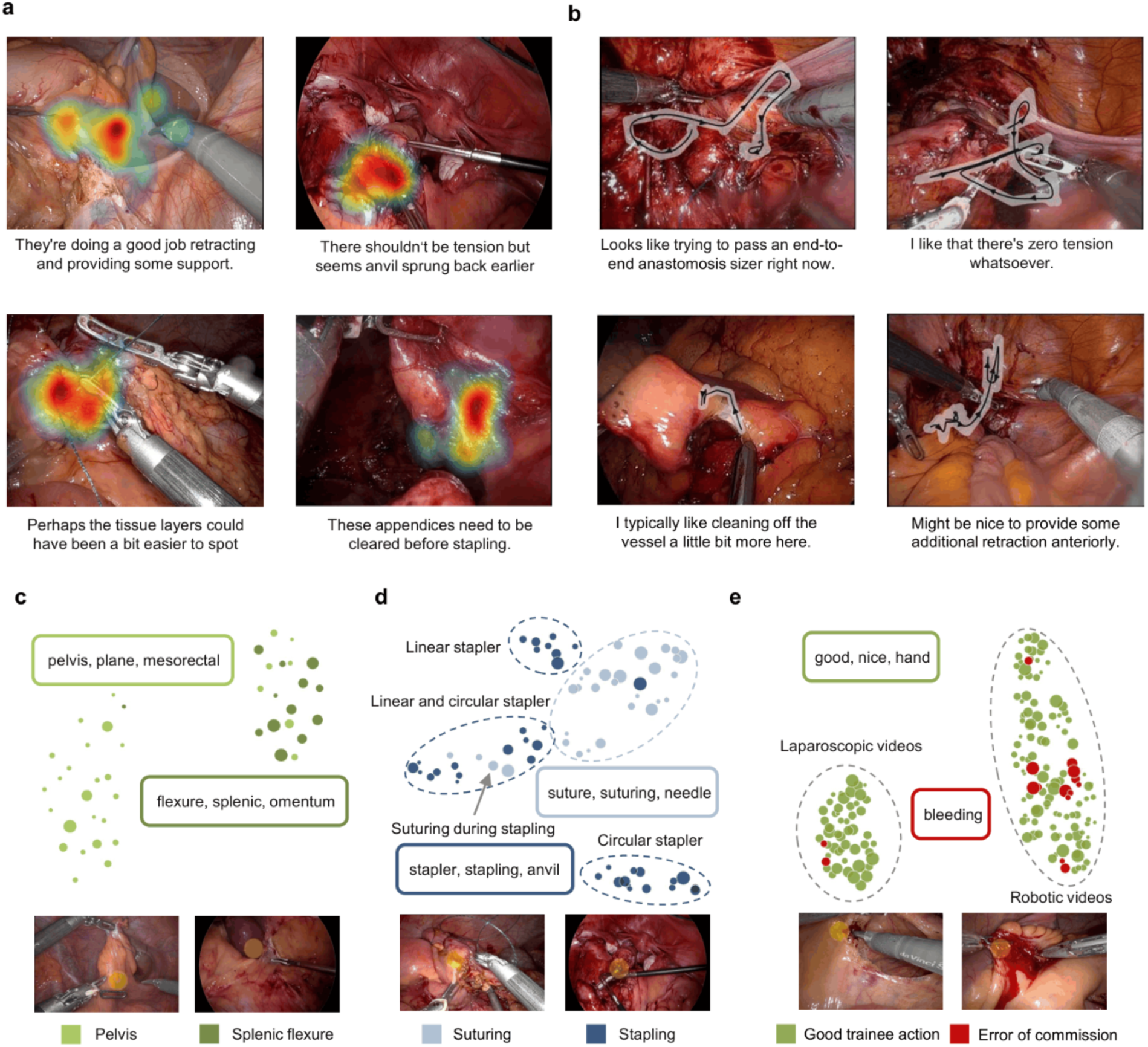
Multimodal capture of surgical commentary and spatial annotation, and visual-embedding structure of implicit annotations. a, b, Representative frames illustrating the simultaneous capture of verbal commentary and spatial attention data during video review. In each panel, the reviewer’s spoken feedback (shown below representative scene images) was recorded alongside their spatial attention, aggregated over the full duration of that verbal segment. **a,** Eye gaze data, overlaid as a density map indicating reviewer visual attention during feedback. **b,** Cursor tracking data, overlaid as a trajectory tracing where the reviewer pointed while speaking. Together these modalities provide a spatially and temporally aligned record of what the reviewer said and where their visual attention was directed. **c-e,** UMAP projections of visual embeddings extracted from chunk-based **(c, d)** or trigger-based **(e)** surgical video clips. Clips were identified based on chunks that contained mutually exclusive keywords **(c, d)** or had mutually exclusive triggers **(e)**, frames were sampled from the respective chunk-based clips and encoded with a self-supervised vision model (DINOv2). The resulting per-clip embedding was projected to two dimensions with UMAP. Each point represents one clip, coloured by category and sized by the word count of its associated verbal commentary chunk. **c,** Anatomical structures (keywords: “mesorectum”, “mesorectal”, “rectal fascia”, “mesorectal fascia”, “rectal plane”, “mesorectal plane”, “pelvis”, “pelvic” vs. “splenic flexure”). **d,** Surgical actions (“suture”, “suturing”, “stitch”, “sew” vs. “staple”, “stapler”, “stapling”); within stapling, linear and circular staplers form separate sub-clusters. **e,** Feedback triggers (good trainee action vs. error of commission); here, the embedding separates laparoscopic from robotic footage rather than the two trigger classes.

To assess the spatial information encoded in eye gaze trajectories captured during feedback narration, we compared the visual appearance of the video when reviewers narrated distinct anatomy (rectum vs. splenic flexure), distinct technical concepts (suturing vs. stapling), and distinct feedback triggers (good action vs. critical triggers), using UMAP. These embeddings were derived from the video appearance itself, using only clips that passed our eye gaze quality filter (i.e., those in which the reviewer’s gaze remained on the surgical field for a sufficient proportion of the segment), as in our main experiments.

Visual embeddings of these video sections were most separable by anatomical region (Fig. 4c; silhouette score: 0.39) and remained separable by technical concept (Fig. 4d; silhouette score: 0.20), where distinct stapler types (linear vs. circular) further formed their own sub-clusters. In contrast, embeddings for different feedback triggers overlapped substantially (Fig. 4e; silhouette score: -0.10): here the visual signal instead separated robotic from laparoscopic footage, rather than good from erroneous actions. This is consistent with the high variability in appropriate and erroneous surgical actions, as well as the evaluative distinction being a higher-level judgement that is not purely based on visual appearance of a brief period of a single chunk.

Together, these findings indicate that meaningful spatial information can be captured alongside verbal commentary without additional expert effort, and that the two signals jointly encode clinically relevant concepts.

### Usability of implicit surgical video annotation

The implicit annotation method received a mean System Usability Scale (SUS) score of 83.8 ± 8.6 across 15 reviewers (Fig. 5), indicating usability in the "excellent" range.^25,26^ Per-item responses showed broad agreement on ease of use and learnability.

**Fig. 5.**
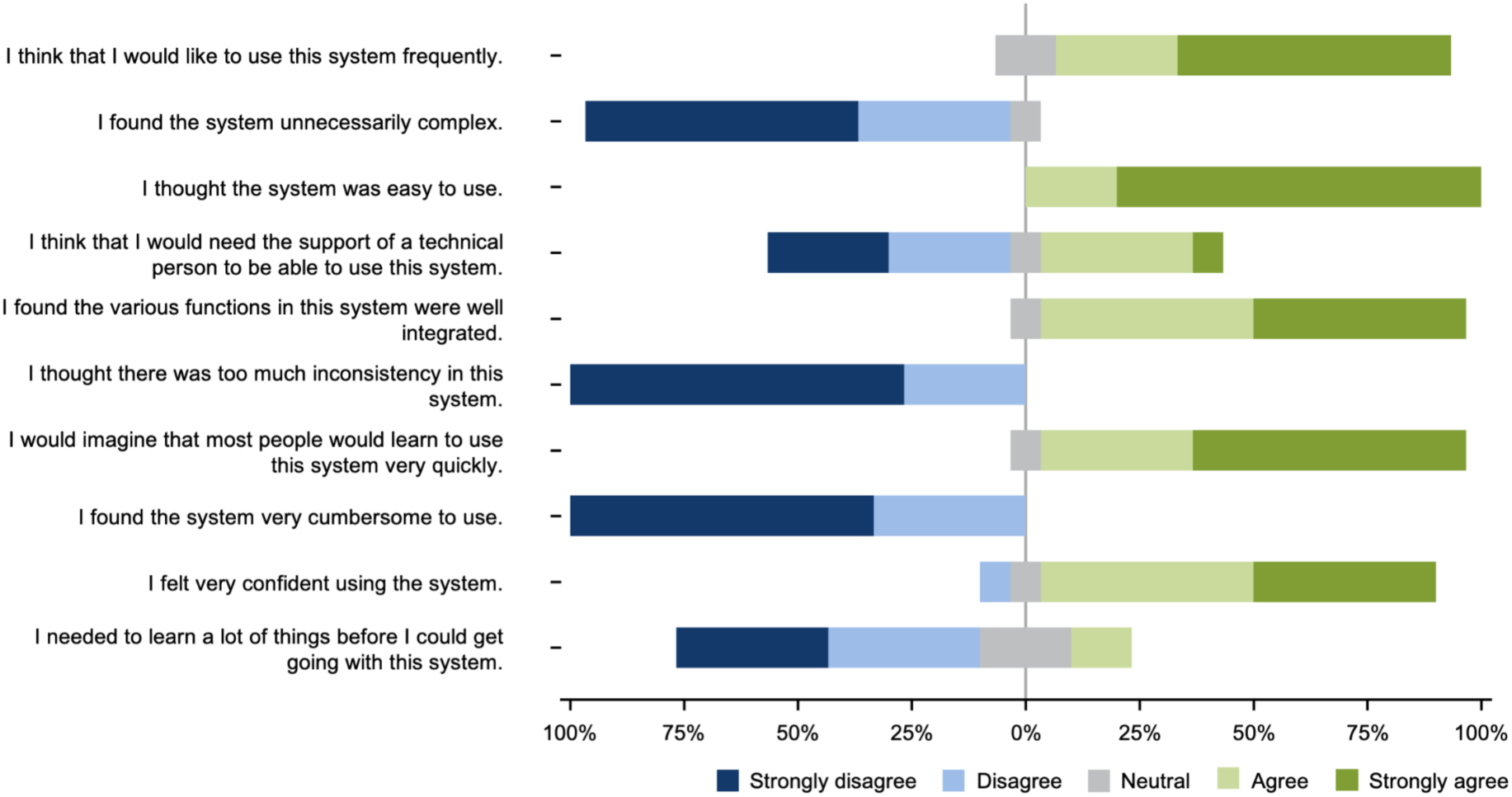
Usability evaluation results for LLM-augmented implicit surgical video annotation. Likert-scale responses to the 10-item System Usability Scale (SUS) administered to all reviewers (n = 15) after completion of the annotation task. Bars are diverging-stacked: bars to the left of the centre line represent disagreement (dark blue = strongly disagree, light blue = disagree), grey represents neutral, and bars to the right represent agreement (light green = agree, dark green = strongly agree).

## Discussion

The objective of this research was to develop a scalable method to contextualize surgical video data with complex expert reasoning signals. We show that spoken language and visual attention can be captured and converted into structured, machine-actionable records of intraoperative behaviours. Across two contrasting and clinically relevant annotation tasks, video-based peer feedback on full-length colorectal procedures and structured CVS assessment in laparoscopic cholecystectomy, implicit annotation captured accurate labels that facilitate downstream data science analyses and spatiotemporal insights at a level of continuity, contextualization, and granularity that existing surgical data science methods do not offer. Implicit annotation captured these complex annotations at several times real-time video speed, and surgeons rated the method as highly usable. These characteristics position implicit annotation to contribute to addressing annotated data scarcity, a persistent barrier to progress in surgical data science.^12^

Existing approaches to surgical video annotation largely map visual or kinematic signals onto a predefined target. Most publicly available surgical video datasets were manually annotated with comparably simple, descriptive label.^15^ Generating labels conveying more complex, context-dependent judgments in an automated or semi-automated manner has proven challenging due to their high variability and context dependence, as well as the scarcity of publicly available respective data that could be used for training or finetuning computational tools, especially foundation models. Existing surgical data science-based systems for video understanding, for example those that track instrument motion to output a binary skill classification or a continuous skill score in laparoscopic cholecystectomy, can perform well on the rating they are trained to reproduce but compress expert judgement into a single label and do not reflect context-dependent reasoning processes.^27^ More recent systems decode multiple elements of intraoperative activity, from surgical subphases to gestures and skill, and some can surface post-hoc explanations that highlight the video frames or motion features most responsible for a given prediction. Yet these explanations describe the model’s own output rather than the operating team’s reasoning, and they remain confined to predefined taxonomies of activity^28,29^. A parallel line of work in surgery constructs video-language supervision from signals that happen to accompany public videos, most often narrated lectures, yielding large question-answer corpora but inheriting the selectivity of teaching narration, which emphasises didactic points over routine operative steps.^30^ Implicit annotation takes the expert’s own verbal reasoning as the primary signal, retaining why an action was judged as it was and where in the procedure, and thereby preserves the semantic content by design. It is scalable, captures finer-grained judgements than descriptive labels, and adapts to different annotation tasks simply by adjusting the language-model prompt.

We evaluated gaze tracking with a high-frequency eye tracking system and cursor tracking on a screen as two distinct modes of enriching verbal patterns and video with spatial signals. Each of these modes has distinct strengths that map onto discrete intended settings. Cursor tracking does not require specific hardware and needs no calibration, making it more readily scalable to high-volume retrospective review settings, at the cost of lower temporal density and a dependence on active reviewer engagement. Eye tracking yields dense, passively acquired attention data at no additional cost to the reviewer, and suits environments where voluntary movement of a cursor would be impractical, yet dedicated hardware and calibration are generally feasible, including potential intraoperative real-time data collection during procedures. Eye tracking has previously been used extensively across surgical training and assessment, including in the operating room^31^, and gaze captured intraoperatively has recently been shown to support automated inference of clinical roles, surgical phase and team communication.^32^ Overall, we regard cursor capture to be the more practical method for large-scale post-hoc annotation of existing, uncontextualized surgical video data, whereas eye tracking may be preferable where rich spatial signal needs to be collected hands-free, e.g., capture during live surgery.

Reviewers rated the method as highly usable (mean SUS 83.8). While this is in the upper range of established benchmarks for interpreting SUS scores ^25,26^, 6 of 15 reviewers still felt they would need help from a technical person to use it. This suggests a main barrier lies in initial setup and training rather than the review task itself. Guided onboarding processes and a task-related rater training curriculum would likely contribute to eased adoption, usability, and, ultimately, interpretability of annotations.

This study has limitations. First, we validated the method on two applications in a representative, yet limited sample of 10 full-length colorectal surgeries and publicly available cholecystectomy clips from 32 cases. As such, this study provides validation of the annotation method, however, it cannot establish broad insights across procedure types, institutions and levels of reviewer experience, and it similarly cannot link video content to clinically relevant endpoints, including complications. Second, reviewers received instructions that were related to use of the method and devices, but received no task-specific training or calibration beyond the task prompt and criterion definitions, and therefore applied their own internalised standards without worked examples or prior alignment on how each criterion should be judged. We acknowledge that structured rater training would likely have made the commentary more homogeneous and, during implicit analysis, easier to chunk, classify, and use in downstream analyses. We accepted this limitation deliberately, because it reproduces natural review practice and tests the method against the expected variability it must tolerate to be useful and robust. The observed performance should accordingly be read as a lower bound, reflecting unrehearsed expert speech and unaligned individual standards, which the method reproduced expert labels despite. Third, 64 of 349 chunks were excluded from the spatial analysis because the quality of the eye tracking signal over the corresponding segment was insufficient. This is a known failure mode of eye tracking that arises when calibration drifts during a session^33^; it affects only the spatial component and leaves the semantic results unchanged, and it is addressable through review protocols that include repeated calibration checks during a session, or through head-mounted eye tracking systems that tolerate calibration drift better. Taken together, this work shows how expert surgical reasoning can be captured as it naturally occurs during video review, and that these signals can be made machine-actionable at scale, addressing a persistent barrier in surgical data science and broadening the applications of surgical data science.

## Outlook

Surgical reasoning has so far been difficult to capture and convert to structured labels at scale. Implicit surgical video annotation drastically lowers the cost of building annotated surgical video datasets, and expands the range of information types that such datasets can be contextualized with. Current datasets are annotated almost exclusively with narrowly defined, descriptive labels, which support models that recognise what is happening but not why a given action was appropriate or how it might have been done differently. By capturing evaluative and counterfactual reasoning anchored in time and in the operative field, implicit annotation opens applications that descriptive labels cannot support. It could underpin systems that surface alternative technical approaches during an operation. It could link specific intraoperative behaviours to downstream clinical outcomes such as complications and turn operative video into a traceable, searchable record of surgical decision-making. They also provide a basis for benchmarking surgical reasoning rather than surgical description, which is what current vision-language models are evaluated on. Overall, we anticipate that implicit annotation will reduce the barrier that data scarcity currently presents to progress in surgical data science.

## Methods

### Implicit Review Method Implementation

#### Software

The implicit annotation method was implemented as a browser-based application requiring no software installation beyond a standard web browser. During a session, it displayed the procedure recording alongside a task-specific prompt and, where applicable, on-screen assessment controls (per-criterion selectors for the CVS task). For full-length procedure review, the playback speed was adjustable from 0.5× to 16× (default 5×), and reviewers could pause, scrub, and replay any segment. The application recorded the continuous audio stream together with a log of the video position displayed at each moment, and, where collected, gaze or cursor coordinates. This yielded an audio-time-to-video-time mapping so that every verbal expression could be anchored to its position in the source video, including across variable-speed and re-watched segments. Audio was subsequently transcribed with a domain-prompted automatic speech recognition model (Whisper, OpenAI, San Francisco, CA, USA) and processed through the LLM-based chunking and classification pipeline that composes the implicit annotation method (Supplementary Information 3-9).

#### Hardware

Review sessions were run on a standard desktop workstation with a 27-inch monitor (1920×1080) in a quiet room. Audio was captured with standard headphones (Apple AirPods Pro, Apple Inc., Cupertino, CA, USA). Where spatial annotation was collected, on-screen gaze was recorded with a desktop-mounted Gazepoint GP3 HD eye tracker (Gazepoint, Vancouver, BC, Canada). The web-based configuration required no specialised hardware beyond a standard computer, a web browser, a standard computer mouse, and a paired audio device, with spatial context captured through cursor position rather than eye tracking.

### Study Design

#### Ethical Approval

All data collection was approved by the Purdue University Institutional Review Board (IRB #2025-00000221). Before participation, informed consent was obtained. No identifying information was stored alongside the review data, audio recordings were deleted after transcription had been verified, and all data were held on Purdue University’s secure file transfer protocol.

#### Surgeon Participant Selection

Surgeons spanning a range of training levels, from residents to attending surgeons, and subspecialty backgrounds were recruited at Indiana University School of Medicine. Eligible participants were attending surgeons and residents with experience in general or colorectal surgery. For the open-review task, nine surgeons each reviewed the full-length colorectal recordings. For the CVS task, eight surgeons each reviewed a subset of clips, with every clip reviewed independently by two surgeons. Before review, each participant completed a structured entry form capturing participant identifier, gender, years of surgical experience, surgical specialty, spectacle wear, and procedure-specific experience level (observer, assistant, or primary surgeon).

#### Video Review Process

Each review session was conducted with a single reviewer seated alone in a quiet room at the workstation described above. Reviewers completed a standardized microphone quality check to ensure adequate signal acquisition, followed by standardized, task-specific instructions via a pre-recorded video and standard 9-point Gazepoint calibration for eye tracking. During the session, the procedure recording was displayed alongside the task prompt, and reviewers narrated their assessment continuously while controlling playback, slowing down or replaying critical or ambiguous segments as needed. Playback speed was adjustable from 0.5× to 16×, with a default of 5× for full-length procedures. Task-specific prompts and the corresponding reference standards are described in the task sections below (open-ended peer feedback and CVS assessment). As a proof of principle, we additionally implemented a fully web-based version of the platform in which spatial context was captured through cursor tracking rather than eye tracking, removing the need for dedicated hardware.

#### Usability Evaluation

After completing the annotation task(s), reviewers rated the usability of the implicit review platform using the System Usability Scale (SUS), a validated 10-item questionnaire with responses on a five-point Likert scale. Item scores were combined into an overall usability score ranging from 0 to 100 following the standard SUS scoring procedure.^25^ The questionnaire was administered once per reviewer; surgeons who participated in both the CVS and open-review tasks completed the SUS a single time and were counted once. A total of 15 distinct reviewers completed the questionnaire.

### Implicit video-based feedback of full-length colorectal surgery videos

#### Dataset

Full-length recordings of minimally invasive colorectal procedures were obtained from an in-house dataset curated prospectively at Indiana University Health University Hospital as part of clinical routine care. Patients who underwent colorectal surgery between 10/28/2025 and 03/26/2026 and for whom a complete full-length intraoperative video recording was available were eligible for inclusion. A total of 10 videos were included, comprising sigmoid colectomy, rectopexy, bowel resection, low anterior rectal resection, abdominoperineal resection and right hemicolectomy, performed by both laparoscopic and robotic approaches, with durations ranging from 69 to 160 minutes. Case-level metadata are provided in Supplementary Information 11.

#### Review process

Each recording was independently reviewed by three surgeons of varying years of experience (Supplementary Information 12). Reviewers were instructed to provide video-based feedback on aspects of surgical quality relevant to patient outcomes, and were prompted: *"Review the entire surgical procedure and comment on surgical quality, defined as intraoperative behaviours that impact patient outcomes. If you were giving feedback while this surgeon was operating, what would you tell them? Examples include general categories of technical execution, decision-making/judgement, safety/critical moments, or specific things to work on."* Reviewers controlled playback throughout (0.5-16×, default 5×) and could pause and replay any segment to examine critical or ambiguous moments in greater detail.

#### Chunking

Reviewer commentary was transcribed and segmented into discrete, semantically coherent spans ("chunks"), each of which was subsequently classified. The full chunking protocol, worked examples and benchmarking are provided in Supplementary Information 3-6. Chunking ground truth for the study cases was established by independent manual segmentation of each transcript by two medical students (R.P.B. and K.K.) following the chunking protocol (Supplementary Information 4), with disagreements arbitrated by a physician with training in general surgery (F.R.K.). Annotators were provided with 7 worked examples (Supplementary Information 5), chunked by consensus among three research team members (two graduate researchers [Z.Z. and M.I.Q.] and one physician with training in general surgery [F.R.K.]).

To select the automated chunking model, four lightweight LLMs (Gemini 3 Flash, Qwen 3 Flash, GPT-5 mini, and Claude Sonnet 4.6) were benchmarked in a few-shot configuration on the 7 consensus-chunked development transcripts, using mean cosine similarity to the human ground truth as the selection metric (Supplementary Information 3 and 6). The best-performing model (Claude Sonnet 4.6) was integrated into the pipeline. Automated chunking was then evaluated against the chunking ground truth of the 30 annotated study transcripts, with the protocol and the 7 consensus-chunked transcripts supplied as few-shot examples. Agreement, both model versus human and inter-rater agreement, was quantified as the mean cosine similarity over optimally matched chunk pairs (BGE-small embeddings, L2-normalised, Hungarian one-to-one matching; Supplementary Information 6 and 10).

#### Classification

Each chunk was first classified as noise or codable according to whether it contained surgical content. Each codable chunk was then assigned one or more observation labels (anatomical, procedural, technical, external viewpoint or counterfactual perspective) and at most one mutually exclusive trigger label (error of commission, error of omission, warning or good trainee action), following a scheme adapted from the validated surgical-feedback taxonomy of Wong et al.^23^ (Supplementary Information 7 and 8).

Classification ground truth was established by independent manual labelling of every chunk produced by the automated chunking step by two medical students (R.P.B. and K.K.) following the classification protocol, with disagreements arbitrated by a physician with training in general surgery (F.R.K.). Ground-truth annotators were additionally provided with 100 classified chunks, drawn at random from the 7 example case transcripts and labelled by consensus among three research-team members (two graduate researchers [Z.Z and M.I.Q.] and one physician with training in general surgery [F.R.K.]), as worked examples (Supplementary Information 9).

To select the classification model, four reasoning-capable language models were benchmarked on all chunks from the 30 ground-truth-annotated procedures in a many-shot configuration, with the classification protocol and the 100 consensus-labelled chunks supplied as examples. Models were compared on per-category accuracy and Cohen’s κ for both observation and trigger labels (Supplementary Information 13). The best-performing model (Claude Opus 4.8, Supplementary Information 13) was used for all classification results reported in the main text.

### Implicit video-based annotation of CVS criteria

#### Dataset

Laparoscopic cholecystectomy clips (n = 32) were drawn from the publicly available SAGES Critical View of Safety Challenge dataset^34^. Because each of the three CVS criteria can be graded in binary fashion, there are 8 possible score combinations. We sampled four clips per combination, using the ground truth provided in the public dataset to ensure even coverage across all eight combinations. The mapping between clip identifiers and source files is provided in Supplementary Information 14.

#### Review process

A total of 32 laparoscopic cholecystectomy clips were independently reviewed by two surgeons each, which resulted in 64 commentaries from eight surgeons with varying years of experience. (Supplementary Information 14 and 15). Each clip was 90 seconds long, but reviewers could adjust the replay speed and speak for as long as needed. For each clip, reviewers were prompted: "*Watch each clip and evaluate it based on the three CVS criteria below. Provide verbal commentary on whether each criterion is Achieved or Not Achieved, and explain your reasoning. Make sure your commentary addresses all three criteria."* The three criteria were defined as C1, two-structure view (two tubular structures visible, connected to the gallbladder); C2, hepatocystic triangle (cleared of fat and fibrous tissue); and C3, cystic plate view (lower third separated from the liver bed). In addition to their verbal commentary, reviewers assigned an explicit rating to each criterion at the end of the clip through on-screen selection, which served as the reference standard.

#### Chunking

Because CVS commentary is short and highly structured, reviewer transcripts were segmented sentence-wise, with each sentence treated as one chunk. All other transcription and preprocessing steps were identical to the open-review task (Supplementary Information 3-6).

#### Classification

For each clip, sentences relevant to the CVS were assigned to the corresponding criterion (C1, isolation of the cystic duct and cystic artery; C2, clearance of the hepatocystic triangle; C3, exposure of the cystic plate), while sentences without criterion-relevant content were labelled as noise. From the criteria-related commentary, the best-performing language model (Gemini 3.1 Pro), selected based on the highest Cohen’s κ, then inferred a per-criterion rating for each clip based on the comparison of four models’ performance on this task (Supplementary Information 16). The full prompt, comprising the criterion definitions and the rating instruction, is reproduced in the review process above. These implicit, commentary-derived ratings were compared against the explicit per-criterion ratings that reviewers recorded through the on-screen controls during the same session and which served as the reference standard.

## Statistical analysis

Analyses were performed in Python 3.11 using NumPy, pandas, SciPy, statsmodels and scikit-learn.

Chunk length is reported as the median and interquartile range (IQR) in words, because the distribution of chunk lengths is right-skewed. Content-category co-occurrence was summarised, for each observation category, as the proportion of chunks carrying that label that also carried at least one other observation label. Discussion time per CVS criterion is reported as the mean ± s.d. in seconds.

Chunking agreement was quantified as the mean cosine similarity over optimally matched chunk pairs, using BGE-small embeddings, L2 normalisation and Hungarian one-to-one matching, and aggregated across transcripts as a mean ± s.d. weighted by the number of ground-truth chunks per transcript. Model-versus-human and human-versus-human agreement were computed identically (Supplementary Information 6 and 10).

Agreement between the automated pipeline and the arbitrated human ground truth was quantified with Cohen’s κ. For the colorectal feedback task, to account for the clustering of chunks within videos, κ was computed separately for each video and aggregated across videos as a chunk-weighted mean ± s.d., weighting each video by its number of chunks. Observation labels were treated as independent binary present/absent classifications per category, and trigger labels as a single multiclass variable over the five levels (four triggers plus no-trigger). Videos for which κ was undefined for a given label, that is, with no positive instances in either the model output or the ground truth, were excluded from that label’s aggregate, and the number of contributing videos is reported. For the CVS task, agreement between the model-inferred per-criterion ratings and the explicit reviewer ratings was quantified with Cohen’s κ for each criterion, reported as the mean ± s.d.

Chunk embeddings were projected into two dimensions with UMAP for visualisation; in all cases the projection is unsupervised, and labels were used only to colour the resulting points and did not inform the embedding or the projection. For the colorectal feedback task, chunks were embedded with the Gemini embedding model (gemini-embedding-001, 3,072 dimensions, clustering task type) and projected with UMAP (n_neighbors = 50, min_dist = 0.3, cosine metric, random_state = 42). For the CVS task, chunks were embedded with BGE-small (BAAI/bge-small-en-v1.5) and projected with UMAP (n_neighbors = 50, min_dist = 0.45, spread = 1.2, cosine metric, random_state = 42).

Cluster separation in the 2-D UMAP layouts was quantified with the silhouette score. For single-label projections (CVS criteria, Fig. 3b; visual-embedding clips, Fig. 4c–e), a standard multi-class silhouette score was computed directly over the category labels. For the multi-label colorectal observation projection (Fig. 2c), where a chunk may carry more than one observation category and the silhouette score is not directly defined, a one-vs-rest silhouette was computed per category, with each chunk counted in every category it carried, and summarised as a single value by averaging across categories weighted by category size.

Because each clip contributes paired measurements across CVS criteria, pairwise differences in discussion time between criteria were tested with the Wilcoxon signed-rank test, with P < 0.05 considered significant.

## Data and code availability

Source data and code are available from the corresponding author upon reasonable request. The large language models used for chunking and classification are publicly available commercial models named in the Methods.

## Acknowledgements

Funding

S.M. is funded by the National Institute on Aging (K23 AG071945). F.R.K. receives support from the German Federal Ministry for Research, Technology and Space BMFTR (GRAIL, 01ZU2505), the German Cancer Aid (AI-TME, 70116966), the German Cancer Research Center (CoBot 2.0), the Jung Foundation for Science and Research (Jung Career Advancement Award), the Central Indiana Corporate Partnership AnalytiXIN Initiative, and the Indiana Clinical and Translational Sciences Institute funded, in part, by the National Institutes of Health, National Center for Advancing Translational Sciences, Clinical and Translational Sciences Award (UM1TR004402). The content is solely the responsibility of the authors and does not necessarily represent the official views of the National Institutes of Health.

## Author contributions

Z.Z. and F.R.K. conceptualized this study. Z.Z., M.I.Q., R.R., M.B., and F.R.K. designed the methodology, implemented code and supporting algorithms, optimized and validated the method. E.M.A., K.T.E., M.J.G., S.K.H., B.K.H., B.W.R., T.S., J.A.W., D.S., K.Y.B., S.M., and F.R.K. provided clinical data and implicit annotations. Z.Z., M.I.Q., R.R., M.B., and F.R.K. collected and curated data and R.P.B., K.K., and F.R.K. annotated ground truth labels. Z.Z., M.I.Q., and F.R.K. conducted formal analyses, prepared visualizations, and wrote the initial draft of the manuscript. K.Y.B., S.M., and F.R.K. provided resources and funding support, and F.R.K. provided overall project management and oversight. All authors read, revised, and approved the final manuscript version.

