## Supplementary Information for "Large language model-augmented implicit surgical video review"

### Table of Contents

Supplementary Information 1: Data Collection Methodology 1
Supplementary Information 2: Data Analysis Methodology 3
Supplementary Information 3: Chunking Methodology 4
Supplementary Information 4: Chunking Protocol and Prompt 5
Supplementary Information 5: Examples for Chunking 6
Supplementary Information 6: LLM Benchmarking for Chunking 12
Supplementary Information 7: Adaptation of Validated Classification for Postoperative Feedback 13
Supplementary Information 8: Classification Protocol and Prompt 14
Supplementary Information 9: Examples for Classification 16
Supplementary Information 10: Methodology for Interrater Agreement Calculation 19
Supplementary Information 11: Video Metadata for Video-Based Feedback 20
Supplementary Information 12: Annotator Metadata for Video-Based Feedback 21
Supplementary Information 13: LLM Benchmarking for Classification 22
Supplementary Information 14: Data Sources for Implicit CVS Annotation 24
Supplementary Information 15: Annotator Metadata for Implicit CVS Annotation 25
Supplementary Information 16: LLM Benchmarking for Implicit CVS Annotation 26
References 27

### Supplementary Information 1: Data Collection Methodology

**Ethics and setting.** All data collection was approved by the Purdue University Institutional Review Board (IRB #2025-00000221). Reviewing surgeons provided written informed consent before participation through an on-screen consent step that specified the data to be captured (verbal commentary, on-screen gaze position, assessment responses and basic demographic information), the voluntary nature of participation, and the right to withdraw at any time. No identifying information was stored alongside the review data. Audio recordings were deleted after transcription had been verified.

Each review session was conducted with a single reviewer seated alone in a quiet room, wearing a supplied headset with microphone (AirPods Pro, Apple, Inc., Cupertino, CA, United States) for audio capture and, where applicable, positioned in front of a desktop-mounted eye tracking unit (Gazepoint GP3 HD, Gazepoint Research Inc., Vancouver, BC, Canada). Before any review, microphone signal quality was verified through a short standardised audio check (volume, duration and dynamic-range thresholds), and reviewers re-ran the check until acceptable signal was confirmed. Demographic and experience information was collected through a structured entry form (participant identifier, gender, years of surgical experience, surgical specialty, spectacle wear, and procedure-specific experience level); the resulting reviewer characteristics are summarised in Supplementary Information 12 (video-based feedback) and Supplementary Information 15 (CVS).

**Video sources.** For the video-based review task, 10 full-length minimally invasive colorectal procedure recordings were obtained from Indiana University School of Medicine. Case-level metadata (procedure type, surgical approach, duration and resolution) are listed in Supplementary Information 11. For the structured task, laparoscopic cholecystectomy clips were drawn from the publicly available SAGES Critical View of Safety dataset; the correspondence between clip identifiers and the original source files is provided in Supplementary Information 14.

**Review system.** The review system was implemented as a browser-based application requiring no specialised software beyond a standard web browser and a paired audio capture device. During a session, the procedure recording was displayed alongside the task-specific instruction, and the reviewer narrated continuously while controlling playback. Playback speed was adjustable from 0.5× to 16×, with a default of 5× for full-length procedures; reviewers could pause, scrub and replay any segment. The application recorded the continuous audio stream together with synchronised playback metadata, so that every point in the audio could be mapped back to its corresponding position in the source video.

**Review tasks and prompts.** Each reviewer completed two task types, each preceded by a task-specific instruction and an explicit consent-to-record step. In the CVS task, reviewers narrated their assessment while assigning explicit per-criterion ratings (criterion 1, isolation of the cystic duct and cystic artery; criterion 2, clearance of the hepatocystic triangle; criterion 3, exposure of the cystic plate) through on-screen controls. In the video-based review task, reviewers narrated free-form commentary on surgical quality, covering technical execution, intraoperative decision-making, safety-relevant moments, and aspects to improve, without a pre-specified taxonomy. The verbatim instruction text presented for each task is reproduced below. For the CVS task, reviewers were instructed: "Watch each clip and evaluate it based on the three CVS criteria below. Provide verbal commentary on whether each criterion is Achieved or Not Achieved, and explain your reasoning. Make sure your commentary addresses all three criteria. C1, Two-Structure View: two tubular structures visible, connected to gallbladder. C2, Hepatocystic Triangle: cleared of fat and fibrous tissue. C3, Cystic Plate View: lower third separated from liver bed." For the video-based review task, reviewers were instructed: "Review the entire surgical procedure and comment on surgical quality, defined as intraoperative behaviours that impact patient outcomes. If you were giving feedback while this surgeon was operating, what would you tell them? Examples include general categories of technical execution, decision-making and judgement, safety and critical moments, or specific things to work on. You must watch the entire video; you may adjust playback speed or skip less relevant sections, but please ensure you review the full procedure before completing your evaluation."

**Prospective reference set.** In addition to the review sessions above, 7 annotations were collected for method development purposes, with cursor tracking for spatial contextualization instead of eye tracking. The resulting annotations were used as few-shot examples for the automated chunking and classification steps (Supplementary Information 3–9); they were not included in any reported evaluation result.

### Supplementary Information 2: Data Analysis Methodology

**Handling of replayed and variable-speed segments.** Because reviewers traversed low-complexity segments quickly and slowed down for critical or ambiguous moments, audio time and video time were not linearly related within a session. To preserve anchoring, the system maintained a continuous audio-time to video-time mapping that accounted for pauses, scrubs, speed changes and re-watched segments. When a segment was replayed, commentary was anchored to the video position under review at the moment of speaking rather than to elapsed session time, so that repeated passes over the same anatomy resolved to the same underlying video interval.

**Systematic prompt construction.** Prompts for the automated steps were assembled through a structured prompt builder that composed each prompt from fixed components: a task definition, the relevant protocol (identical to the protocol given to human annotators; Supplementary Information 4 and 8), a fixed set of few-shot examples, and the input transcript. Holding the protocol and example set constant across reviewers and cases ensured that differences in output reflected the input commentary rather than prompt variation.

**Selection of the chunking model.** Four lightweight language models were compared for the chunking step (Supplementary Information 6). Model selection used soft cosine similarity against ground-truth chunk boundaries on a single held-out reference case (CASE-008); the selected model was then re-evaluated on the full set of ground-truth-annotated transcripts (Supplementary Information 6).

**Selection of the classification model.** Four reasoning-capable language models were compared for the classification step. Each was prompted with 100 ground-truth-annotated examples drawn from the 7 prospectively annotated cases, and evaluated on 30 ground-truth case transcripts; selection used classification accuracy and Cohen's $\kappa$ for both trigger and observation labels (Supplementary Information 13). Performance of the final configuration is reported in the main text.

**Supplementary Information 3: Chunking Methodology**

Chunking is the step that segments continuous reviewer commentary into discrete spans, each of which subsequently receives at least one observation label or is marked as noise. Chunking was performed by an instruction-following language model run in a few-shot configuration, with the chunking protocol (Supplementary Information 4) supplied verbatim together with worked examples.

Four candidate models were compared: Gemini 3 Flash, Qwen 3 Flash, GPT-5 mini and Claude Sonnet 4.6. Decoding was left at each model's recommended configuration rather than forced to a fixed temperature, because the reasoning-capable models used here either omit or override a temperature parameter: light reasoning was enabled where supported (Gemini 3 thinking level "low"; Claude adaptive thinking at "low" effort), temperature was left at the provider default for Gemini and Claude and is not accepted by the GPT-5 family, and the output-token budget was set generously (32,768–64,000 tokens) so that reasoning tokens did not truncate the structured output. Identical prompts and worked examples were used across all four models, so that differences in output reflected the model rather than the prompt.

Of the 7 prospectively annotated example transcripts, one representative case (CASE-008) was held out as a test case, and the remaining 6 (CASE-002, CASE-005, CASE-011, CASE-012, CASE-018 and CASE-019) were supplied as few-shot demonstrations during model comparison. The model achieving the highest mean cosine similarity to the human ground-truth chunking on the held-out case (Supplementary Information 6) was selected. The selected model was then re-configured to use all 7 annotated example cases as few-shot demonstrations and applied to produce the final chunking of the 30 annotated study cases.

**Supplementary Information 4: Chunking Protocol and Prompt**

The same chunking protocol was supplied to the human annotators and to the best-performing language model, so that automated and manual chunking followed identical rules. The complete prompt comprised the chunking protocol reproduced below, and the 7 worked few-shot examples rendered as gold standard chunks of raw transcripts (Supplementary Information 5). The model was instructed to return a JSON array in which each element is one chunk, carrying the chunk text and a noise flag, and to preserve the input text verbatim. Every output was checked for lexical preservation: the concatenated chunk text was required to reproduce the source transcript, so that paraphrasing or omission would be detected rather than silently accepted.

**Chunking Protocol for Surgical Review Commentary**

Split each reviewer's continuous commentary into chunks. A chunk is the basic span that later gets at least one observation label (or is marked noise), and may carry one trigger type. A chunk is one continuous semantic span encompassing a single theme or topic. It is often one sentence, but can occasionally span several sentences or a part of a long sentence (split when the theme changes). Every chunk is either codable (has surgical content) or noise (no content, kept as its own chunk rather than deleted). A codable chunk may contain one or multiple observations of trainee actions, anatomic structures, and/or technical or procedural details, and belongs to at most one trigger type.

Start a new chunk when the theme changes, i.e. the main idea, focus, or evaluative stance shifts. Concretely: the action or step changes ("…dissecting. Now they're stapling."); the context moves to a new, unrelated structure, region, technique, or procedural detail (colon → mesorectum); the evaluative stance changes notably (praise → error or warning); or there is a clear discourse break ("okay, moving on") or a content-to-non-content transition. As a default, if in doubt, separate chunks at sentence ends. A theme change inside one long sentence is itself enough to split, without needing a sentence boundary: "good exposure of the colon, but watch the ureter it is not displayed well" → 2 chunks.

Keep in one chunk when following sentences continue the same theme ("They are oversewing this lesion. They are doing a figure of eight."); when fillers sit inside a meaningful sentence ("They are oversewing this lesion. Okay, good. They are doing a figure of eight."); or when a reason or qualifier follows a claim ("Nice plane, staying between the layers.").

**Noise chunks.** A standalone span that is entirely non-content becomes its own noise chunk, for example "Okay." between two sentences, "oh ah yeah there.", or "the uis."; meta-comments such as "can I change the speed" or "let me scrub forward" are also noise. Test: if dropping the span loses nothing about the surgery, it is noise.

**Two situations in one sentence.** Different themes are split into separate chunks; the same theme carrying multiple feedback types stays as one chunk.

**Examples**

| **Chunks** | **Decision** |
|---|---|
| "…redundancy… A stretchy redundant mesentery. In the pelvis." | 1 chunk (continuation) |
| "um, so they're, yeah, mobilizing the colon." | 1 codable chunk (fillers stay) |
| "can I change the speed of the video" | 1 noise chunk |
| "They just finished dissection. Now they're stapling." | 2 chunks (action change) |
| "They are staying in the plane and use less traction" | 1 chunk |

#### Supplementary Information 5: Examples for Chunking

This section provides the worked few-shot examples used to guide the chunking step, illustrating how continuous reviewer commentary is segmented into semantically coherent chunks according to the chunking protocol.

**CASE-002**

So now we're doing a hand assist. So there's a hand assist.
Procedure and patient's thin.
And the hand is looking at the transverse colon.
And moving the small bowel a little bit.
Oh, I see.
So they're holding up the IMB.
And seeing the ureter with Firefly.
Now looking at the appendix.
Seeing the ureter.
So the hand, I expect this is maybe a total collecting or something.
So hand assist starting at the right.
Using energy to mobilize the colon laterally.
Some adhesions of the flexure.
So the hand keeps getting in the way of the camera.
Good. Good vibes with the energy.
Okay.
So the hand is looking at the ureter.
And the hand is looking at the colon. Okay. So the hand is looking at the colon.
Okay.
Good vibes with the energy device.
And good use of the hand to provide retraction.
I'm going to go back to 5x now.
So looks like we're probably doing a total colectomy here.
Looks like the surgeon is pointing out some things to the assistant.
The instruments aren't moving.
Looks like maybe the hand came out and went back in again.
The surgeon is pointing out to the assistant where to go. And the surgeon is doing some finger dissection to move the assistant along in the right plane.
A little bit of bleeding.
Dividing the gastrocolic momentum.
Looks like this one's to be a total colectomy.
Nice use of the hand to facilitate the dissection.
Hand came out and went back in again I think.
It looks like the surgeon is assisting. Okay. Good. The surgeon is assisting him or herself now with a hand and a grasp.
And then directing the assistant where to go.
They've decided they want to preserve the momentum so they corrected. And now preserving some momentum and coming right along.
The transverse colon.
Very successful and efficient right now.
So the surgeon is using a grasper and the hand to guide the assistant with the energy device across the whole transverse colon very nicely.
And now the lesser sac is pretty wide open which helps. Or maybe they're just making it look easy.
So now at the osmotic flexor still approaching it medial to lateral and not medial to lateral but from the transverse colon side.
And nice. Very nice.
I'm guessing this is the total colectomy.
They haven't taken any mesentery yet. They're just doing mobilization.
Using a little bit of a shrivel to look close to the bowel which makes me think this is probably all coming out.
Good use of the hand to facilitate the conception. Preparing each bite by sweeping.
The camera person is a little confused now.
And continuing down the descent colon.
And down the lower abdomen.
The hands getting in the way more as usual.
So they're using the firefly intermittently to see where the ureter is now.
So now back up to the transverse colon.
I think they're going to start taking the mesentery. Actually they're going to do a little more mobilization of the ascending laterally.
So they probably just don't have quite enough to take the mesentery. And holding on to the shadow.
Now that you've got the mesentery working very efficiently,
using the full length of the energy device, and double burning as needed.

Patient's very thin, so you can actually see all the individual vessels.
I usually take the mesentery in segments as I do the mobilization, but they've done this very efficiently, doing it as separate steps.
So I've zoomed around, taken all the mesentery nicely.
And back at the acem colon, probably getting ready to divide the Ti here.
Still using the hand to get effect.
Taking the mesentery right along the bowel.
I'm wondering if this is a colitis patient. Or do they want to not take the iliacolic.
Just completing the mesenteric division at the Ti.
Very nice.
A little more mobilization at the base of the Ti.
And the procedure is done.

**CASE-005**

This appears to be a cystoscopy.
I'm going to skip over it.
It appears to be a direct opti-view technique.
I went through the momentum, but I didn't see any issues.
I'm going to skip over it.
Looks like since it's a sigmoid colectomy,
place the cap and I'm going to slow it back down.
I didn't have any issues with the port placement. All the ports are up.
All the instruments were placed under direct visualization, which is good.
It's a sigmoid cancer, but looks like starting with a lateral to medial mobilization. I'm curious why the lateral to medial mobilization.
To start with, it's not wrong.
I think the frequent ureter checks earlier were good.
This is the IMA. Looks like kind of a large IMA. That the IMA was taken at the appropriate location.
Already confirmed that the ureter is down.
See that there's the vein. Hasn't taken the vein. Not sure if planning on taking it there.
I think they're in a good plane.
Doing fairly frequent ureter checks to make sure the ureter is in the right location.
Really seem to be getting to the upper rectum essentially.
I agree with the angle that taking the mesentery.
See where the tumor is. Clearly this tattoo is fairly distal to where the actual cancer is.
I thought the stapling went well. It's nice that you were able to get it all in one fire. I feel like sometimes that can be difficult to do robotically.
This patient obviously has fairly redundant colon. Doesn't look like reach is going to be an issue. You can tell trying to thin out the mesentery and the epiploid fat to get the help out of the healthy clear colon.
Looks like getting ready to check perfusion to the proximal transection line. Yeah I can see the ICG now.
I agree with the proximal transection point.
I can see this must have been when they're introducing the anvil.
Okay.
I do wonder if the staple line is a little close to where the EEA anvil is. I don't know if the goal is to come across that staple line or to totally exclude it like a side to end. I guess we'll see here in a minute. Okay. Looks like side to end.
Okay.
I just notice had to put decrease the trim down in Berg in order to get the fluid in the pelvis which is nice.
Now as you see the tumor over here so nice distal margin.
Looks like placing grass burr and then just going to extract so I think the majority of the case is done.
So I think overall the quality of the dissection is good.
The only I don't want to say issue but the only concern is I'm not sure why it took down those lateral attachments to then do a medial to lateral mobilization. Seems like from a retraction standpoint it would have just left those medial or excuse me those lateral attachments intact. It could have helped with the medial to lateral mobilization of the mesentery.
Otherwise again a good ligation. No tension.
Blood supply was confirmed.
Leak test was confirmed.
So good operation overall.

**CASE-008** (held-out case for chunking benchmarking, see Supplementary Information 6)

Looks like it's getting in.
Placing the drill curves.
Now, elevating colon.
And then we did the appropriate exposure.
And then we're going to use the assist port to help.
Taking a little bit more lateral attachments.
Looks like doing more of a lateral dissection first.
Looks like it's the lower attachments that are causing the issue.

You can see the ureter.
Still working and gliding down towards the left one.
We're going to put the lumbar out of the way a little bit more.
Now taking the attachments, that's good.
Taking the lumbar out of the pelvis.
Trying to get into the pre-sacral plane. We put in a good plane.
The lumbar is still in the way. We're going to potentially flip it over to get it out of the way.
Now we're going to work on the lateral. The lateral is sort of coming down a little posterior.
Now it looks like taking the vessel.
I'm going to expose the IMA ID to identify the origin of the IMA.
Looks like it's going to urinate a little bit more as well.
Looking for the ureter.
Looks like it's trying to identify the correct plane.
Getting a little bit in the mesentary. Worked to be a little deeper.
Now looking for the ureter. Trying to get it to move. There we go.
Now we're going to work on the IMA to identify the structures.
It doesn't appear that the plane is quite correct. Maybe a little deep.
Looking like a better plane.
Coming through the mesentary again without clearly identifying the origin of the IMA.
I'd like to see better exposure between I and V. Okay. I'm going to see if I can get the I and V is coming through to help identify the artery better.
Looks like trying to clean up the artery.
Still like to get some of that small bowel, particularly to the left side of the screen, out of the way to expose it better.
Looks like it's burning.
Little better flanger.
A little bit of a strange interest above the lining up that needs to come down.
There we go.
The ureter in the background.
Okay.
And the correct plane.
Starting to see some of that vascular plane. Be helpful to open that side. I see it starting to come up this side.
Again, more help with the exposure.
Still trying to open the side more.
More help to go down.
Looking at the anterior, I would still recommend taking the side wall, the lateral stock attachment down a little bit more to find the anterior dissection plane.
Starting a new plane in the anterior.
Okay.
The ureter plane is here.
So, we recommend taking the lateral attachments, which will help get around to the anterior, will help more with exposure.
There, now starting to take these lateral attachments.
Okay.
Some are more posterior to left anterior like they are right there.
There's the plane. That's good.
Okay.
I like starting from the backside. I think it's helpful to start from the backside and follow that dissection around. So, I do like the general pushing up of the ureter action to try to identify the plane.
Looks good here posteriorly. Continue this way.
Also, the lateral attachments. I can see the tension right there. That's nice clean. Continue that dissection up.
It's helpful still to come on this left side.
There you go.
You can see the tension.
Okay.
It would be helpful to release some of the lateral attachments from the anterior side on the left.
Hopefully they're going there now.
Good. That's a good dissection there. I like this dissection now. Okay. There you go. I like that move.
Looks like it's starting to see barrier area rectum.
I'll put the backside section up.
Okay.
Great turn.
Okay.
I'm going to divide the mesentery up to the colon where we'll divide the colon.
Okay.
Looks like very close to having the mesentery clean off the colon where you'll be able to staple across it. And get the staple out all the way across. Sometimes I like to hold a little pressure so it doesn't slide out, but it looks like it did well there.

**CASE-011**

So far I think nice controlled movements, good retraction with the left hand here.
Looks like we're getting the lateral attachments down in the spacial there at prior surgery or I guess it looks like probably prior surgery with some adhesive disease over here.
I don't think I've seen the ureter yet but it seems like we just took the pedicle.
Seems like we're still trying to look for the ureter in that retroperitoneal tissue.
Doing a nice job of gently dissecting out that retroperitoneal tissue,
making sure things are safe before taking bites along here.
I like the retraction here in the dissection making sure that we're identifying any important structures before taking bites with the ligature.
I think I was mistaken before. Well, I'm five times speed,
but it looks like this is the pedicle knot would be additional tissue that they took in the mesentery previously.
Looks like we're designing our distal transection margin.
Looks like we're under the rectum here,
we're circumferentially separating the fat from the rectum.
Looks like we have a sizer or something to help facilitate to make sure that we can get up to the transection margin for the anastomosis.
I like this additional retraction.
I was having a hard time seeing where the rectal edges were. So holding it in this manner makes it a little bit easier to dissect and make sure not cutting into the rectum. I think that was a good move.
I like this tunneling, making sure the rectum is up and the meso rectum is down.
I don't love the hematoma there on the rectum. Probably okay but I would just keep an eye on it before we do any of the anastomosis.
Good retraction here, good separation from the retroperitoneal tissue, taking those lateral attachments.
Looks like the hematoma stays the same size. Less concerned about doing an anastomosis with that now, making sure that it's not expanding.
Looks like we maybe mobilized the omentum for having a nice tension free anastomosis.
I like that we're separating some of this to bring that up.
Good traction there.
Nice position with the stapler.
Looks like a nice tension free anastomosis.
I believe the mesentery is flat, I guess I haven't seen that fully with the view that we've had so far.
We're doing a leak test. Looks like we're doing a leak test now.
Oh, that was a good case.
Good efficiency of motion,
good retraction,
very precise movements,
good dissection,
nice and careful making sure you're identifying important structures before doing any transection.

**CASE-012**

So ** [age removed for deidentification] year old patient.
We need to show them what we need for dissection.
You can see the difference.
I never say that dramatically, but obviously, not much to comment on here.
Experienced hands clearly working to take down the adhesions.
Maybe work a little bit wider here if possible. Although in general, this looks just like that. This is expert. This is an expert clearly.
This is really nice dissection.
I typically like cleaning off the vessel a little bit more here, but it's a personal preference.
Looks like the omentum is being left behind here. My preference for a case like this one would be to include the omentum at least for the portion. Let me see the portion that's attached to the colon that's gonna be resected. So I'll split the omentum up towards the border of the transverse colon. And then you get into the space above and then include all of the omentum with the specimen. That's proximal to the tumor.
Generally very efficient movement.
I'm really watching the maneuver with some crossing of the instruments that for a case where you're only doing it without an assistant, completely understandable.
Really nice dissection,
don't really have much in the way of formative comments overall.
Very, very nice exemplary.
And this portion of the procedure looks like the resection of the pelvic mass.

**CASE-018**

This is a cystoscopy, so not relevant for our quality.
Now we're in the abdomen,
preparing to start.
So this is a hand assist operation.

I can see the surgeon's hand in the gel port.
The surgeon is using a hand assist approach to work on the IMA pedicle, which is interesting because I don't typically do that. I usually do this part straight lap because I find that my hand gets in my way, but this surgeon seems to be doing it very efficiently.
The surgeon is taking the vascular pedicle with the energy device
and there's a little bit of bleeding from the pedicle, but it's quickly controlled by the surgeon.
So the surgeon is very efficiently opening the lesser sac and separating the omentum from the transverse colon.
Very nice use of the hand assist technique.
The energy device seems to be failing to seal some of the vessels throughout the case. I wonder if a double burning technique might be useful. This is a different energy device than I use, so... I don't know.
It might be useful to have a raytex sponge in the surgeon's hand to be able to quickly dab the areas of bleeding.
Whoever was holding the camera was shaking a lot earlier in the case and now they're doing a lot better.
So there's still some bleeding up in the omentum,
just hearkening back to the energy device and whether it's really effectively sealing some of these vessels. If it was me I would I would put a ray tech in there and just uh work on that bleeding area. I don't think the surgeon thinks it's bleeding and it may not be, so that comment might not be very useful.
The field is a little more bloody than I like.
And I think it's just from doing a lot of blunt dissection, which is very nice and efficient and has been in the right plane the whole time. (technique-related)
But I wonder if this patient may be particularly prone to little hemorrhages from the dissection. (more general, patient-related)
The video has paused now.
I don't know what's happening. (disfluency)
I wonder if they're getting ready for the extraction. (procedural_meta)
Okay, now we're back to it.
ligating the base of the descending colon mesentery.
I think the surgeon is feeling the colon to decide where the proximal resection margin should be, which is a nice use of the hand.
Yeah, I think that's exactly what's going on.
Thinking about doing some Firefly here, I think.
I don't love how they're just sitting around waiting for something to happen right now. It's probably the anesthesia provider giving the ICG intravenously, but next time perhaps they could give it a little bit sooner.
All right, now we've got green.
The surgeon is sort of mopping up a little bit of blood that's in the field, which I think is awesome.
that pain, putting the omentum back where it belongs. Agree with that.
Taking a look for any bleeding I think.
I'm actually doing something with the omentum.
I don't know what's going on here.
I think maybe creating a little bit of a flap.
There's some more bleeding here from the energy device, which is not sealing the vessels very well. I would suggest double burning when there's a visible vessel.
Now we're kind of right on the stomach wall, which I think can take a joke.
Now I'm just holding a little pressure.
So it's still bleeding and there's some hemostatic material in there. But I think that's not going to work because it looks like a vessel is bleeding. So I would suggest to focus in on that area and try to dissect out the vessel and just carefully ligate it.
Cleaning the camera.

**CASE-019**

Noting that you already have a cap in place and have a trocar through there.
See the other trocars.
The colostomy takedown, I'm assuming that the cap is at the colostomy site.
And fairly minimal adhesive disease, which is nice.
I'm assuming at this point, trying to find the colostomy stump.
Looks like probably won't need to do much proximal colon mobilization.
Just getting the ports into place.
instruments into place at least.
Looks like the patient has a lot of adhesions at the prior colovesical fistula site. Hopefully this doesn't cause too much issue.
Well that looks like some granulation.
I think I saw earlier that an assist port was placed but I don't see that the assistant is actively helping.
Might be nice to have them provide some additional retraction anteriorly.
Now the assistance here obviously with dissection.
I'm not sure if this patient has ureteral stents since it's a Hartman's takedown, but but I would say, I would assume that if you had stents, would have also used ICG in the stents and would have highlighted the ureters by now, just as a quick check, but I'll keep watching and see if that happens at some point later.
Certainly kind of in this area is where it would be most concerned, but it seems that doing just a fine job of getting away from that area without really being in the danger zone.
I'm assuming you were probably the primary surgeon who did the original colectomy just based on the fact that there's not a lot of sigmoid mesentery. I feel like a emergency general surgeon would have left a bunch of thick mez there.

You're doing a good job of just kind of taking time opening up this plane trying to get the rectal stump really isolated and away from all the inflammation.
Looks like at this point you're trying to highlight where the ureter would be since probably don't have stents.
Looks like probably trying to pass an EEA sizer right now.
Or performing a colonoscopy.
tell since that video is not included. Yeah, clearly something. Okay.
So this person appears to only have one prolene. Maybe I missed the other one, which is not an issue. But sometimes it's nice to have one on either side so you know both sides.
Again, you can see the rectum pretty well.
And it looks like when you put the EEA up, it's gonna be just fine on away from everything else. I think this is always the most painful part of an EEA with a robotic platform. There's nothing that grabs it very well. I will say the medium clip applier or the Cobra Grasper, which is like the cardiac probe Grasper is actually really good at grabbing the anvil. I think the clip applier is probably easier thing to get and is more commonly used, but I think the Maryland and the bipolar just are too small and don't hold the grip very well. (all about grabbing the anvil)
I like that there's zero tension whatsoever. nice and smoothly.
Yossa, with having two smaller Graspers it may might be hard to occlude the colon for the leak test but we'll see what you use. Using a bigger Grasper like the tips up fenestrated or the small grasping retractor just has a longer longer tips and can usually occlude the entire bowel to use for this part.
It looks like you're not using the robotic bed because it's not paired.
So it's hard to tell how much Trendelenburg the patient is in, but obviously it can be another nice adjustment to do a little less Trendelenburg.
You can see the flexible scope coming up. Obviously it looks good.
I don't see any any pressure, any bubbles, nothing.
So pretty well constructed anastomosis.
that loop of small bowel.
All right, I think that went pretty well overall.

### Supplementary Information 6: LLM Benchmarking for Chunking

**Agreement metric.** Chunking agreement was quantified as the mean cosine similarity between optimally matched chunk pairs. For a given pair of segmentations (model versus human, or human versus human), each chunk was embedded with the BAAI General Embedding model (BGE-small; embeddings were L2-normalised, so that the cosine similarity between two chunks equals their dot product). For each case we formed the similarity matrix between the two sets of chunks and computed an optimal one-to-one matching using the Hungarian algorithm (linear sum assignment), which maximises the total similarity of the matched pairs; for cases with unequal chunk counts, the matching pairs the smaller of the two counts. The reported agreement is the mean cosine similarity over the matched pairs. Mean cosine similarity was the sole agreement metric used for chunking.

**Model selection (held-out case).** Mean cosine similarity on the held-out case (CASE-008) is shown in Supplementary Information 6. Claude Sonnet 4.6 achieved the highest agreement and was selected for the final chunking.

| Metric | Gemini 3 Flash | Qwen 3 Flash | GPT-5 mini | Claude Sonnet 4.6 |
|---|---|---|---|---|
| Mean cosine similarity | 0.95 | 0.95 | 0.96 | 0.98 |

**Final chunking agreement (selected model).** For the final chunking, agreement between the selected model (Claude Sonnet 4.6) and the arbitrated human ground truth was computed for each of the 30 study transcripts separately. For each transcript, chunks from the two segmentations were matched one-to-one with the Hungarian algorithm and the mean cosine similarity over matched pairs was taken as that transcript's agreement. The reported value is the mean ± s.d. across the 30 transcript-wise means, with each transcript weighted by its number of ground-truth chunks. This yielded a chunk-weighted mean cosine similarity of 0.951 ± 0.013 across the 30 transcripts.

**Supplementary Information 7: Adaptation of Validated Classification for Postoperative Feedback**

The classification scheme was adapted from the validated live-feedback taxonomy of Wong et al. for use in postoperative narration over a recording.[1] Three changes follow from the postoperative setting (as compared to the intraoperative setting used for the original taxonomy): First, the trainer and trainee response axes of the original system were removed, because no trainee is present during review to respond, request clarification, or be taken over from. Second, the trainee-question trigger was removed, because no live trainee asks questions during review. Third, feedback classes were modified to better reflect the nuances of postoperative commentary: Visual Aid, Praise and Criticism were removed as classes, and External Viewpoint, Counterfactual Perspective, and Noise were added based on commonly observed patterns in feedback narrated post-hoc. A detailed description of the final taxonomy used in this study is available in Supplementary Information 8.

The adapted scheme therefore retains two axes: an observation axis describing what the reviewer is observing (anatomical, procedural, technical, external viewpoint or counterfactual perspective), and a trigger axis describing the reviewer's evaluative stance toward an observed trainee action (error of commission, error of omission, warning, or good trainee action). Each codable chunk carries at least one observation label and at most one trigger; chunks may carry multiple observation labels simultaneously.

### Supplementary Information 8: Classification Protocol and Prompt

You will be presented with written chunks of surgical feedback, which are derived from narrated feedback based on a replayed video and can encompass a part of a sentence, one sentence, or multiple sentences on the same theme or topic. Every chunk is either codable (has surgical content) or noise (no content, kept as its own chunk, not deleted). A codable chunk can contain one or multiple observations of trainee actions, anatomic structures, and procedural or technical details. Each chunk belongs to at most one Trigger type.

#### Observation (at least one required per codable chunk)

A codable chunk must carry one or more of the following. A chunk may carry several at once, for example anatomic and technical observations together.

1. **Anatomical.** Reference to an anatomic structure, plane, or landmark in the patient's abdomen. Example: "stay in the correct plane," "between the two fascial layers."
2. **Procedural.** Timing, sequence, or choice of a surgical step or approach. The primary goal of this feedback is to optimize and understand the overall strategy and sequence of the case. Example: "starting with a lateral to medial mobilization," "switch to the left side now."
3. **Technical.** Execution of a discrete, intentional, active task, including exposure, instrument handling, traction, and visualization. This primarily describes which task is carried out and how it is carried out. Example: "placed under direct visualization," "buzz it."
4. **External Viewpoint.** Description of personal preferences or assumptions related to the feedback, often based on personal experience and information gathered outside of this case. Examples: "I sometimes use the third arm for improving exposure in this step", "This is often difficult in obese patients".
5. **Counterfactual Perspective.** Philosophical expressions or elaborations about a different (earlier or later) timepoint that are hypothetical. Examples: "I wonder if it would have been better to do the stapling earlier".

#### Trigger (none or one per chunk, only when an evaluative stance is present)

A Trigger is assigned only when the chunk contains the reviewer's evaluative stance toward an observed trainee action. It is possible for a chunk to contain only observations and no Trigger, but a chunk cannot contain multiple triggers, as they are mutually exclusive.

1. **Error of commission.** The reviewer identifies an incorrect action that produced a definable mistake. Example: "never use the traumatic grasper like that there, it caused tissue injury."
2. **Error of omission.** The reviewer notes a needed action done partially or not at all. Example: "be helpful to open that side," "you need to go more laterally."
3. **Warning.** The reviewer flags suboptimal or risky behavior with no definable mistake yet. Example: "careful, they are getting close to the ureter here."
4. **Good trainee action.** The reviewer affirms an acceptable or correct action. Example: "which is good," "nice plane."

#### Noise definition

A standalone span that is entirely non-content becomes its own noise chunk (examples: noise / meta-comment): No content at all ("Hm.", "Okay.") Meta-comments about the data collection process. Extreme transcription errors rendering contextualization and classification impossible.

### Examples

| Chunk | Content | Trigger |
|---|---|---|
| "All the instruments were placed under direct visualization, which is good." | Technical | Good trainee action |
| "It's a sigmoid cancer, but looks like starting with a lateral to medial mobilization. I'm curious why the lateral to medial mobilization. To start with, it's not wrong." | Procedural | Good trainee action |
| "Starting to see some of that vascular plane." | Anatomical | none |
| "Be helpful to open that side, this is not good." | Technical | Error of omission |
| "Um, so they're, yeah, mobilizing the colon." | Procedural, Anatomical | none |
| "Can I change the speed of the video." | noise | none |

### Supplementary Information 9: Examples for Classification

| Chunk | Observation | | | | | Trigger | | | | Noise |
|---|---|---|---|---|---|---|---|---|---|---|
| | A | P | T | E | C | Error of com. | Error of om. | War-ning | Good action | |
| I went through the omentum, but I didn't see any issues. | Yes | No | No | No | No | No | No | No | Yes | No |
| This is the IMA. Looks like kind of a large IMA. That the IMA was taken at the appropriate location. | Yes | No | No | No | No | No | No | No | Yes | No |
| Already confirmed that the ureter is down. | Yes | Yes | No | No | No | No | No | No | No | No |
| See that there's the vein. Hasn't taken the vein. Not sure if planning on taking it there. | Yes | No | No | No | No | No | Yes | No | No | No |
| Doing fairly frequent ureter checks to make sure the ureter is in the right location. | Yes | No | Yes | No | No | No | No | No | Yes | No |
| Really seem to be getting to the upper rectum essentially. | Yes | No | No | No | No | No | No | No | No | No |
| I agree with the angle that taking the mesentery. | Yes | No | Yes | No | No | No | No | No | Yes | No |
| See where the tumor is. Clearly this tattoo is fairly distal to where the actual cancer is. | Yes | No | No | No | No | No | No | No | No | No |
| This patient obviously has fairly redundant colon. Doesn't look like reach is going to be an issue. You can tell trying to thin out the mesentery and the epiploid fat to get the help out of the healthy clear colon. | Yes | No | No | Yes | No | No | No | No | No | No |
| I do wonder if the staple line is a little close to where the EEA anvil is. I don't know if the goal is to come across that staple line or to totally exclude it like a side to end. I guess we'll see here in a minute. Okay. Looks like side to end. | Yes | No | Yes | No | Yes | No | No | Yes | No | No |
| The only I don't want to say issue but the only concern is I'm not sure why it took down those lateral attachments to then do a medial to lateral mobilization. Seems like from a retraction standpoint it would have just left those medial or excuse me those lateral attachments intact. It could have helped with the medial to lateral mobilization of the mesentery. | Yes | Yes | Yes | Yes | Yes | No | No | Yes | No | No |
| So they're holding up the IMB. | Yes | No | No | No | No | No | No | No | No | No |
| Now looking at the appendix. | Yes | No | No | No | No | No | No | No | No | No |
| Seeing the ureter. | Yes | No | No | No | No | No | No | No | No | No |
| Using energy to mobilize the colon laterally. | Yes | No | Yes | No | No | No | No | No | No | No |
| So the hand keeps getting in the way of the camera. | No | No | Yes | No | No | No | No | Yes | No | No |
| So the hand is looking at the ureter. | Yes | No | Yes | No | No | No | No | No | No | No |
| And the hand is looking at the colon. Okay. So the hand is looking at the colon. | Yes | No | Yes | No | No | No | No | No | No | No |
| And good use of the hand to provide retraction. | No | No | Yes | No | No | No | No | No | Yes | No |
| I'm going to go back to 5x now. | No | No | No | No | No | No | No | No | No | Yes |
| So looks like we're probably doing a total colectomy here. | No | Yes | No | No | No | No | No | No | No | No |
| Looks like the surgeon is pointing out some things to the assistant. | No | No | No | No | No | No | No | No | No | Yes |
| The surgeon is pointing out to the assistant where to go. And the surgeon is doing some finger dissection to move the assistant along in the right plane. | Yes | No | Yes | No | No | No | No | No | No | No |
| Hand came out and went back in again I think. | No | No | Yes | No | No | No | No | No | No | No |
| It looks like the surgeon is assisting. Okay. Good. The surgeon is assisting him or herself now with a hand and a grasp. | No | No | Yes | No | No | No | No | No | Yes | No |
| And then directing the assistant where to go. | No | No | Yes | No | No | No | No | No | No | No |
| The transverse colon. | Yes | No | No | No | No | No | No | No | No | No |
| And now the lesser sac is pretty wide open which helps. Or maybe they're just making it look easy. | Yes | No | No | No | No | No | No | No | Yes | No |
| So now at the osmotic flexor still approaching it medial to lateral and not medial to lateral but from the transverse colon side. | Yes | Yes | No | No | No | No | No | No | No | No |
| Using a little bit of a shrivel to look close to the bowel which makes me think this is probably all coming out. | Yes | Yes | Yes | No | No | No | No | No | No | No |
| The camera person is a little confused now. | No | No | Yes | No | No | No | No | Yes | No | No |
| Now that you've got the mesentery working very efficiently, | Yes | No | No | No | No | No | No | No | Yes | No |
| Patient's very thin, so you can actually see all the individual vessels. | Yes | No | No | No | No | No | No | No | No | No |

| Chunk | Observation | | | | | Trigger | | | | Noise |
|---|---|---|---|---|---|---|---|---|---|---|
| | A | P | T | E | C | Error of com. | Error of om. | War-ning | Good action | |
| I usually take the mesentery in segments as I do the mobilization, but they've done this very efficiently, doing it as separate steps. | Yes | Yes | No | Yes | No | No | No | No | Yes | No |
| So I've zoomed around, taken all the mesentery nicely. | Yes | No | No | No | No | No | No | No | Yes | No |
| And back at the acem colon, probably getting ready to divide the Ti here. | Yes | No | No | Yes | No | No | No | No | No | No |
| Still using the hand to get effect. | No | No | Yes | No | No | No | No | No | No | No |
| I'm wondering if this is a colitis patient. Or do they want to not take the iliacolic. | Yes | No | No | Yes | Yes | No | No | Yes | No | No |
| Just completing the mesenteric division at the Ti. | Yes | Yes | Yes | No | No | No | No | No | No | No |
| Looks like it's getting in. | No | No | No | No | No | No | No | No | No | Yes |
| Still working and gliding down towards the left one. | No | No | Yes | No | No | No | No | No | No | No |
| We're going to put the lumbar out of the way a little bit more. | Yes | Yes | Yes | No | No | No | No | No | No | No |
| Now taking the attachments, that's good. | Yes | No | Yes | No | No | No | No | No | Yes | No |
| Trying to get into the pre-sacral plane. We put in a good plane. | Yes | No | Yes | No | No | No | No | No | Yes | No |
| Now we're going to work on the lateral. The lateral is sort of coming down a little posterior. | Yes | Yes | No | No | No | No | No | No | No | No |
| Now it looks like taking the vessel. | Yes | No | No | No | No | No | No | No | No | No |
| I'm going to expose the IMA ID to identify the origin of the IMA. | Yes | Yes | Yes | No | No | No | No | No | No | No |
| Looks like it's going to urinate a little bit more as well. | No | No | No | No | No | No | No | No | No | Yes |
| It doesn't appear that the plane is quite correct. Maybe a little deep. | Yes | No | No | No | No | Yes | No | No | No | No |
| Okay. | No | No | No | No | No | No | No | No | No | Yes |
| And the correct plane. | Yes | No | No | No | No | No | No | No | Yes | No |
| Still trying to open the side more. | Yes | No | Yes | No | No | No | No | No | No | No |
| More help to go down. | No | No | Yes | No | No | No | No | No | No | No |
| Looking at the anterior, I would still recommend taking the side wall, the lateral stock attachment down a little bit more to find the anterior dissection plane. | Yes | Yes | No | Yes | No | No | Yes | No | No | No |
| There, now starting to take these lateral attachments. | Yes | No | Yes | No | No | No | No | No | No | No |
| Also, the lateral attachments. I can see the tension right there. That's nice clean. Continue that dissection up. | Yes | Yes | Yes | No | No | No | No | No | Yes | No |
| It's helpful still to come on this left side. | Yes | No | Yes | No | No | No | No | No | No | No |
| I like that there's zero tension whatsoever. nice and smoothly. | No | No | Yes | No | No | No | No | No | Yes | No |
| Hopefully they're going there now. | No | No | Yes | No | Yes | No | No | No | No | No |
| Looks like it's starting to see barrier area rectum. | Yes | No | No | No | No | No | No | No | No | No |
| I'll put the backside section up. | No | No | No | No | No | No | No | No | No | Yes |
| Great turn. | No | No | Yes | No | No | No | No | No | Yes | No |
| Looks like very close to having the mesentery clean off the colon where you'll be able to staple across it. And get the staple out all the way across. Sometimes I like to hold a little pressure so it doesn't slide out, but it looks like it did well there. | Yes | Yes | Yes | Yes | No | No | No | No | Yes | No |
| Looks like we're getting the lateral attachments down in the spacial there at prior surgery or I guess it looks like probably prior surgery with some adhesive disease over here. | Yes | Yes | No | Yes | No | No | No | No | No | No |
| I don't think I've seen the ureter yet but it seems like we just took the pedicle. | Yes | No | Yes | No | No | No | No | Yes | No | No |
| Doing a nice job of gently dissecting out that retroperitoneal tissue, | Yes | No | Yes | No | No | No | No | No | Yes | No |
| making sure things are safe before taking bites along here. | No | No | Yes | No | No | No | No | No | Yes | No |
| I like the retraction here in the dissection making sure that we're identifying any important structures before taking bites with the ligature. | No | No | Yes | No | No | No | No | No | Yes | No |
| Looks like we're designing our distal transection margin. | Yes | Yes | Yes | No | No | No | No | No | No | No |
| Looks like we're under the rectum here, | Yes | No | No | No | No | No | No | No | No | No |
| I don't love the hematoma there on the rectum. Probably okay but I would just keep an eye on it before we do any of the anastomosis. | Yes | Yes | No | Yes | No | No | No | Yes | No | No |
| I believe the mesentery is flat, I guess I haven't seen that fully with the view that we've had so far. | Yes | No | No | No | No | No | No | No | No | No |
| Good efficiency of motion, | No | No | Yes | No | No | No | No | No | Yes | No |

| Chunk | Observation | | | | | Trigger | | | | Noise |
|---|---|---|---|---|---|---|---|---|---|---|
| | A | P | T | E | C | Error of com. | Error of om. | War-ning | Good action | |
| Experienced hands clearly working to take down the adhesions. | Yes | No | Yes | No | No | No | No | No | Yes | No |
| Generally very efficient movement. | No | No | Yes | No | No | No | No | No | Yes | No |
| I'm really watching the maneuver with some crossing of the instruments that for a case where you're only doing it without an assistant, completely understandable. | No | No | Yes | Yes | No | No | No | Yes | No | No |
| Really nice dissection, | No | No | Yes | No | No | No | No | No | Yes | No |
| don't really have much in the way of formative comments overall. | No | No | No | No | No | No | No | No | No | Yes |
| Very, very nice exemplary. | No | No | No | No | No | No | No | No | No | Yes |
| This is a cystoscopy, so not relevant for our quality. | No | Yes | No | No | No | No | No | No | No | No |
| Now we're in the abdomen, | Yes | Yes | No | No | No | No | No | No | No | No |
| preparing to start. | No | Yes | No | No | No | No | No | No | No | No |
| So the surgeon is very efficiently opening the lesser sac and separating the omentum from the transverse colon. | Yes | No | Yes | No | No | No | No | No | Yes | No |
| It might be useful to have a raytex sponge in the surgeon's hand to be able to quickly dab the areas of bleeding. | No | No | Yes | Yes | Yes | No | No | Yes | No | No |
| The video has paused now. | No | No | No | No | No | No | No | No | No | Yes |
| I think the surgeon is feeling the colon to decide where the proximal resection margin should be, which is a nice use of the hand. Yeah, I think that's exactly what's going on. | Yes | Yes | Yes | No | No | No | No | No | Yes | No |
| Taking a look for any bleeding I think. | No | No | Yes | No | No | No | No | No | No | No |
| They are actually doing something with the omentum. | Yes | No | No | No | No | No | No | No | No | No |
| Now I'm just holding a little pressure. | No | No | Yes | No | No | No | No | No | No | No |
| Cleaning the camera. | No | No | Yes | No | No | No | No | No | No | No |
| The colostomy takedown, I'm assuming that the cap is at the colostomy site. | No | Yes | No | Yes | No | No | No | No | No | No |
| Just getting the ports into place. | No | Yes | No | No | No | No | No | No | No | No |
| instruments into place at least. | No | Yes | No | No | No | No | No | No | No | No |
| Looks like the patient has a lot of adhesions at the prior colovesical fistula site. Hopefully this doesn't cause too much issue. | Yes | No | No | No | No | No | No | Yes | No | No |
| Might be nice to have them provide some additional retraction anteriorly. | No | No | Yes | No | Yes | No | No | Yes | No | No |
| tell since that video is not included. Yeah, clearly something. Okay. | No | No | No | No | No | No | No | No | No | Yes |
| And it looks like when you put the EEA up, it's gonna be just fine on away from everything else. I think this is always the most painful part of an EEA with a robotic platform. There's nothing that grabs it very well. I will say the medium clip applier or the Cobra Grasper, which is like the cardiac probe Grasper is actually really good at grabbing the anvil. I think the clip applier is probably easier thing to get and is more commonly used, but I think the Maryland and the bipolar just are too small and don't hold the grip very well. | No | No | Yes | Yes | No | No | No | Yes | No | No |
| Yossa, with having two smaller Graspers it may might be hard to occlude the colon for the leak test but we'll see what you use. Using a bigger Grasper like the tips up fenestrated or the small grasping retractor just has a longer longer tips and can usually occlude the entire bowel to use for this part. | Yes | Yes | Yes | Yes | No | No | No | No | No | No |
| It looks like you're not using the robotic bed because it's not paired. | No | No | Yes | No | No | No | No | No | No | No |
| So it's hard to tell how much Trendelenburg the patient is in, but obviously it can be another nice adjustment to do a little less Trendelenburg. | No | Yes | No | Yes | No | No | No | Yes | No | No |

**Supplementary Information 10: Methodology for Interrater Agreement Calculation**

**Chunking agreement.** Because chunk boundaries are not categorical labels, agreement between two segmentations (model versus human, or human versus human) was quantified as the mean cosine similarity over optimally matched chunk pairs, as described in Supplementary Information 6 (BGE-small embeddings, L2-normalised; Hungarian one-to-one matching; mean cosine over matched pairs). Because the two segmentations of a transcript may contain different numbers of chunks, the Hungarian assignment matches $\min(n_1, n_2)$ pairs. The cosine similarity therefore quantifies semantic agreement on the content captured by both segmentations, while differences in segmentation granularity are reported separately as the chunk-count ratio.

**Aggregation across transcripts.** One cosine similarity was computed for each of the 30 transcripts. These transcript-wise means were then aggregated as a mean ± standard deviation, weighted by the number of chunks in the arbitrated human ground-truth segmentation of that transcript. Because the weights derive from the ground truth rather than from either of the compared segmentations, they are identical across all benchmarked models and across the human–human comparison, so the resulting values are directly comparable. Inter-rater agreement between the two human annotators was computed and aggregated identically.

**Chunking validation (30 cases).** Applying the selected model to all 30 study cases and comparing against the arbitrated human ground truth, the weighted mean cosine similarity was 0.951 ± 0.013 (unweighted mean across transcripts 0.955 ± 0.019; 1,399 model chunks versus 1,450 ground-truth chunks, yielding 1,323 matched pairs).

**Inter-rater agreement (human–human).** As a reference for the achievable agreement on this task, two human annotators independently chunked the same 30 transcripts. Their weighted mean cosine similarity, computed and aggregated identically, was 0.963 ± 0.013. The agreement between the selected model and the arbitrated human ground truth (0.951 ± 0.013) is therefore comparable to the agreement between two independent human annotators on the same transcripts.

**Classification agreement.** For the categorical labels (observation and trigger), agreement between the automated pipeline and the arbitrated human ground truth was quantified using Cohen's κ and overall accuracy (main text, Fig. 2d,e). To account for the clustering of chunks within videos, κ was computed separately for each video and aggregated across videos as a chunk-weighted mean ± s.d., weighting each video by its number of chunks. Observation labels were treated as independent binary present/absent classifications per category, and trigger labels as a single multiclass variable over the five levels (four triggers plus no-trigger).

**Supplementary Information 11: Video Metadata for Video-Based Feedback**

The following table outlines the mapping of individual cases across procedure types, patient demographics, and video specifications.

| Case ID | Procedure Type | Patient age range | Patient sex | Minimally invasive surgery type | Duration | Resolution |
|---|---|---|---|---|---|---|
| P1 | Sigmoid Colectomy | 36-40 years | Female | Laparoscopic | 1:09:11 | 1920x1080 |
| P2 | Rectopexy | 56-60 years | Male | Robotic | 1:26:29 | 1024x768 |
| P3 | Sigmoid Colectomy | 26-30 years | Female | Laparoscopic | 1:27:39 | 1920x1080 |
| P4 | Bowel Resection | 41-45 years | Female | Robotic | 2:15:52 | 1920x1080 |
| P5 | Low Anterior Rectal Resection | 56-60 years | Male | Robotic | 2:40:14 | 1440x1080 |
| P6 | Abdominoperineal Resection | 26-30 years | Male | Robotic | 1:38:45 | 1312x1064 |
| P7 | Right Hemicolectomy | 46-50 years | Female | Robotic | 2:40:02 | 1024x768 |
| P8 | Bowel Resection | 31-35 years | Female | Laparoscopic | 2:12:41 | 1920x1080 |
| P9 | Low Anterior Rectal Resection | 56-60 years | Female | Robotic | 2:31:01 | 1328x1052 |
| P10 | Sigmoid Colostomy | 76-80 years | Female | Laparoscopic | 1:43:36 | 1920x1080 |

**Supplementary Information 12: Annotator Metadata for Video-Based Feedback**

The following table outlines the baseline characteristics and professional experience of the eight independent annotators who provided video-based feedback via implicit video review.

| Annotator ID | Gender | Years of experience | Glasses | Surgical specialty | Colorectal surgery experience |
|---|---|---|---|---|---|
| 1 | Male | 15 | Yes | Colorectal | Primary |
| 2 | Male | 1 | No | General | Observer |
| 3 | Female | 4 | Yes | General | Assistant |
| 4 | Female | 21 | Yes | Colorectal | Primary |
| 5 | Female | 11 | Yes | Colorectal | Primary |
| 6 | Male | 11 | No | Colorectal | Primary |
| 7 | Male | 7 | Yes | Colorectal | Primary |
| 8 | Male | 3 | Yes | General | Assistant |
| 9 | Female | 2 | No | Vascular | Observer |

**Supplementary Information 13: LLM Benchmarking for Classification**

The following table outlines the performance metrics of the evaluated LLMs compared against the manually annotated ground truth for classifying surgical commentary into the observation and trigger taxonomy.

Accuracy and Cohen's κ were computed separately for each surgery and then averaged across surgeries, weighting each surgery by its number of chunks; values are reported as this chunk-weighted mean ± standard deviation. For each category, only surgeries in which that category was present in the ground truth were included, because κ is undefined when a category never occurs. The number of surgeries contributing to each category (n) is given in the column headers.

Observations:

| | **Anatomical (n = 29)** | | **Technical (n = 30)** | | **Procedural (n = 30)** | | **External Viewpoint (n = 28)** | | **Counterfactual perspective (n = 18)** | | **All** | |
|---|---|---|---|---|---|---|---|---|---|---|---|---|
| **LLM** | **Accuracy** | **Kappa** | **Accuracy** | **Kappa** | **Accuracy** | **Kappa** | **Accuracy** | **Kappa** | **Accuracy** | **Kappa** | **Accuracy** | **Kappa** |
| Claude Opus 4.8 | 0.90 ± 0.06 | 0.78 ± 0.13 | 0.84 ± 0.05 | 0.68 ± 0.11 | 0.83 ± 0.06 | 0.56 ± 0.17 | 0.92 ± 0.05 | 0.51 ± 0.26 | 0.95 ± 0.04 | 0.47 ± 0.36 | 0.89 ± 0.03 | 0.71 ± 0.07 |
| GPT-5.5 | 0.92 ± 0.06 | 0.80 ± 0.13 | 0.81 ± 0.05 | 0.61 ± 0.12 | 0.82 ± 0.06 | 0.52 ± 0.20 | 0.91 ± 0.05 | 0.46 ± 0.25 | 0.95 ± 0.05 | 0.51 ± 0.34 | 0.88 ± 0.03 | 0.69 ± 0.08 |
| Gemini 3.1 Pro | 0.90 ± 0.07 | 0.77 ± 0.13 | 0.82 ± 0.06 | 0.64 ± 0.12 | 0.83 ± 0.06 | 0.54 ± 0.18 | 0.91 ± 0.04 | 0.50 ± 0.24 | 0.95 ± 0.04 | 0.58 ± 0.29 | 0.89 ± 0.03 | 0.70 ± 0.06 |
| Qwen3-Max | 0.86 ± 0.08 | 0.68 ± 0.16 | 0.82 ± 0.06 | 0.63 ± 0.12 | 0.80 ± 0.06 | 0.49 ± 0.19 | 0.91 ± 0.04 | 0.54 ± 0.24 | 0.93 ± 0.04 | 0.38 ± 0.24 | 0.87 ± 0.03 | 0.66 ± 0.07 |

Triggers:

| | **Good trainee action (n = 29)** | | **Error of commission (n = 11)** | | **Error of omission (n = 20)** | | **Warning (n = 25)** | | **No trigger (n = 30)** | | **All** | |
|---|---|---|---|---|---|---|---|---|---|---|---|---|
| **LLM** | **Accuracy** | **Kappa** | **Accuracy** | **Kappa** | **Accuracy** | **Kappa** | **Accuracy** | **Kappa** | **Accuracy** | **Kappa** | **Accuracy** | **Kappa** |
| Claude Opus 4.8 | 0.96 ± 0.03 | 0.82 ± 0.16 | 0.97 ± 0.03 | 0.43 ± 0.29 | 0.94 ± 0.05 | 0.47 ± 0.31 | 0.92 ± 0.05 | 0.46 ± 0.32 | 0.89 ± 0.06 | 0.72 ± 0.15 | 0.85 ± 0.08 | 0.68 ± 0.14 |
| GPT-5.5 | 0.95 ± 0.04 | 0.83 ± 0.13 | 0.98 ± 0.03 | 0.50 ± 0.35 | 0.91 ± 0.07 | 0.43 ± 0.28 | 0.93 ± 0.05 | 0.56 ± 0.27 | 0.86 ± 0.07 | 0.67 ± 0.16 | 0.83 ± 0.08 | 0.65 ± 0.16 |
| Gemini 3.1 Pro | 0.95 ± 0.03 | 0.82 ± 0.11 | 0.97 ± 0.03 | 0.40 ± 0.31 | 0.92 ± 0.08 | 0.45 ± 0.31 | 0.92 ± 0.05 | 0.52 ± 0.27 | 0.87 ± 0.07 | 0.69 ± 0.15 | 0.83 ± 0.10 | 0.66 ± 0.15 |
| Qwen3-Max | 0.95 ± 0.03 | 0.80 ± 0.12 | 0.96 ± 0.03 | 0.22 ± 0.30 | 0.94 ± 0.05 | 0.29 ± 0.32 | 0.89 ± 0.07 | 0.40 ± 0.24 | 0.86 ± 0.07 | 0.67 ± 0.16 | 0.82 ± 0.09 | 0.62 ± 0.17 |

Accuracy and Cohen's κ were computed from a single confusion matrix pooled over all chunks, disregarding surgery boundaries and without weighting or averaging across surgeries. No standard deviation is therefore reported.

Observations (overall pooled estimate):

| | **Anatomical** | | **Technical** | | **Procedural** | | **External Viewpoint** | | **Counterfactual perspective** | | **All** | |
|---|---|---|---|---|---|---|---|---|---|---|---|---|
| **LLM** | **Accuracy** | **Kappa** | **Accuracy** | **Kappa** | **Accuracy** | **Kappa** | **Accuracy** | **Kappa** | **Accuracy** | **Kappa** | **Accuracy** | **Kappa** |
| Claude Opus 4.8 | 0.9 | 0.79 | 0.84 | 0.69 | 0.83 | 0.59 | 0.92 | 0.61 | 0.96 | 0.46 | 0.89 | 0.71 |
| GPT-5.5 | 0.92 | 0.82 | 0.81 | 0.63 | 0.82 | 0.56 | 0.91 | 0.54 | 0.95 | 0.46 | 0.88 | 0.7 |
| Gemini 3.1 Pro | 0.9 | 0.78 | 0.82 | 0.65 | 0.83 | 0.58 | .091 | 0.6 | 0.96 | 0.57 | 0.89 | 0.71 |
| Qwen3-Max | 0.86 | 0.7 | 0.82 | 0.64 | 0.8 | 0.54 | 0.91 | 0.63 | 0.94 | 0.37 | 0.87 | 0.66 |

Triggers (overall pooled estimate):

| | **Good trainee action** | | **Error of commission** | | **Error of omission** | | **Warning** | | **No trigger** | | **All** | |
|---|---|---|---|---|---|---|---|---|---|---|---|---|
| **LLM** | **Accuracy** | **Kappa** | **Accuracy** | **Kappa** | **Accuracy** | **Kappa** | **Accuracy** | **Kappa** | **Accuracy** | **Kappa** | **Accuracy** | **Kappa** |
| Claude Opus 4.8 | 0.96 | 0.85 | 0.98 | 0.45 | 0.95 | 0.52 | 0.93 | 0.53 | 0.89 | 0.75 | 0.85 | 0.7 |
| GPT-5.5 | 0.95 | 0.83 | 0.98 | 0.51 | 0.93 | 0.4 | 0.93 | 0.59 | 0.86 | 0.7 | 0.83 | 0.68 |
| Gemini 3.1 Pro | 0.95 | 0.83 | 0.97 | 0.42 | 0.94 | 0.44 | 0.93 | 0.57 | 0.87 | 0.71 | 0.83 | 0.68 |
| Qwen3-Max | 0.95 | 0.81 | 0.98 | 0.33 | 0.95 | 0.33 | 0.89 | 0.44 | 0.86 | 0.69 | 0.82 | 0.63 |

**Supplementary Information 14: Data Sources for Implicit CVS Annotation**

The following table outlines the mapping and origin of individual video segments from the public SAGES CVS Challenge dataset that were reviewed during implicit CVS annotation[2]. Each clip is tracked by their unique identifier linked back to its source video alongside the specific identified CVS annotators who evaluated it.

| Clip ID | SAGES CVS Challenge Dataset Clip ID | Annotated by annotator ID |
|---|---|---|
| 1 | 00467596-8200-449c-8528-d4816ec2f6a2.mp4 | 1, 2 |
| 2 | 01950655-a507-4514-abe5-d13ff593ec4d.mp4 | 3, 4 |
| 3 | 022231f4-37ca-492a-bdc9-cf36b0e37909.mp4 | 1, 2 |
| 4 | 0405e4b0-843a-469d-a235-6a12383bc1db.mp4 | 3, 4 |
| 5 | 04f433b9-bcb8-4753-af4c-d4ca4a701955.mp4 | 1, 2 |
| 6 | 0bce2116-1b33-4b11-a6ac-fe623b0936e8.mp4 | 3, 4 |
| 7 | 0e5b6b69-2ddb-497e-bc25-aad5fe062df8.mp4 | 1, 2 |
| 8 | 1091030d-17d1-4d76-87c0-a917633fc5bf.mp4 | 3, 4 |
| 9 | 16e33e34-9e96-4156-b4ce-7591dcf813da.mp4 | 1, 2 |
| 10 | 18a428a6-9db4-4825-9572-039e4c0b5853.mp4 | 3, 4 |
| 11 | 24ad0a4b-903d-44d3-b74b-c7fb5f906ec0.mp4 | 1, 2 |
| 12 | 280aaeb3-c03c-4fe5-92ba-1e68ac5c2a45.mp4 | 3, 4 |
| 13 | 284ec32e-6b2c-457e-b7f0-0e9517f8f3d5.mp4 | 1, 2 |
| 14 | 2d3be687-240d-4853-bb4a-3ab7fbcc8d9c.mp4 | 3, 4 |
| 15 | 3012e3a3-139c-4e3d-a385-d29420284f6b.mp4 | 1, 2 |
| 16 | 3e037e48-7d37-4397-8a62-bf6926005e84.mp4 | 3, 4 |
| 17 | 42de6aad-1eaf-4a90-9e97-9ddc80fdc32c.mp4 | 5, 6 |
| 18 | 453f5a11-d359-4795-8c40-6132e47a9a87.mp4 | 7, 8 |
| 19 | 52fa51d6-7b5e-44bf-a10c-39bf2022068f.mp4 | 5, 6 |
| 20 | 5eb56c91-26a5-40fc-995d-2694c09b8643.mp4 | 7, 8 |
| 21 | 660a6a01-b2ad-40e1-a709-79c57b1c8fa7.mp4 | 5, 6 |
| 22 | 664a48be-a38f-4bdf-a18d-d902af8da49c.mp4 | 7, 8 |
| 23 | 6722e31c-25a6-41b5-a3ec-ef275919ce49.mp4 | 5, 6 |
| 24 | 7567a428-5fe3-4e2c-a63c-b31f63e684fa.mp4 | 7, 8 |
| 25 | 78a8511a-f204-4e4b-88f2-cf4ab66464f9.mp4 | 5, 6 |
| 26 | 794109cb-4635-414e-954c-2fb499c7036b.mp4 | 7, 8 |
| 27 | 7kibp961-xx4a-i9o6-pshj-ijz9pxc62kjf.mp4 | 5, 6 |
| 28 | 82b01844-2e49-4572-a83f-2b4ddbe56cf9.mp4 | 7, 8 |
| 29 | 8504ff91-785b-4f35-9998-4edd607d48e3.mp4 | 5, 6 |
| 30 | 850e2e68-46a9-4b66-b6f7-3c6de72af7e6.mp4 | 7, 8 |
| 31 | 898c9065-6499-40fa-9142-152ebd5d7227.mp4 | 5, 6 |
| 32 | 93bd2883-8f16-44c5-8588-208bc87c3585.mp4 | 7, 8 |

**Supplementary Information 15: Annotator Metadata for Implicit CVS Annotation**

The following table outlines characteristics of each rater contributing to the implicit CVS annotations analyzed in this work. Annotator characteristics were gathered through a deidentified survey at the end of each annotation session.

| **Annotator ID** | **Gender** | **Years of experience** | **Glasses** | **Surgical specialty** | **Laparoscopic cholecystectomy experience** |
|---|---|---|---|---|---|
| 1 | Female | 4 | Yes | General | Assistant |
| 2 | Female | 2 | No | General | Assistant |
| 3 | Female | 4 | No | General | Assistant |
| 4 | Male | 3 | Yes | General | Assistant |
| 5 | Female | 2 | No | General | Assistant |
| 6 | Male | 3 | Yes | General | Primary |
| 7 | Male | 2 | Yes | General | Assistant |
| 8 | Male | 1 | No | General | Assistant |

### Supplementary Information 16: LLM Benchmarking for Implicit CVS Annotation

| LLM | C1 Accuracy | C1 Kappa | C2 Accuracy | C2 Kappa | C3 Accuracy | C3 Kappa | All Accuracy | All Kappa |
|---|---|---|---|---|---|---|---|---|
| Claude Opus 4.8 | 0.88 ± 0.12 | 0.71 ± 0.23 | 0.70 ± 0.13 | 0.39 ± 0.16 | 0.84 ± 0.15 | 0.68 ± 0.27 | 0.81 ± 0.10 | 0.59 ± 0.18 |
| GPT-5.5 | 0.86 ± 0.12 | 0.68 ± 0.24 | 0.72 ± 0.16 | 0.39 ± 0.27 | 0.88 ± 0.15 | 0.76 ± 0.28 | 0.82 ± 0.08 | 0.61 ± 0.16 |
| Gemini 3.1 Pro | 0.92 ± 0.09 | 0.83 ± 0.18 | 0.78 ± 0.13 | 0.49 ± 0.25 | 0.91 ± 0.11 | 0.81 ± 0.22 | 0.87 ± 0.05 | 0.70 ± 0.09 |
| Qwen3-Max | 0.78 ± 0.15 | 0.50 ± 0.25 | 0.67 ± 0.18 | 0.39 ± 0.22 | 0.84 ± 0.16 | 0.69 ± 0.30 | 0.77 ± 0.13 | 0.53 ± 0.22 |
